# Cerebrospinal fluid clearance impairment captured using stable isotope labeling kinetics (SILK) in normal pressure hydrocephalus

**DOI:** 10.1101/2025.02.11.25322069

**Authors:** CA Leckey, TA Giovannucci, EC Murphy, E Moncur, K Tariq, A Aslanyan, MS Scholl, M Srikrishna, W Coath, S Barker, D Esguerra, A Toma, L Watkins, L Thorne, S Lehmann, J Vialaret, S Wray, RJ Bateman, K Mills, DL Elbert, L Pellegrini, RW Paterson

## Abstract

Normal pressure hydrocephalus is a common cause of gait and cognitive impairment in later life, characterised by accumulation of excessive cerebrospinal fluid (CSF). Clinical improvement can occur following CSF diversion. No biomarkers are available to mechanistically investigate fluid accumulation, support diagnosis or predict response to CSF diversion. We developed a stable isotope labeling kinetics (SILK) method to capture the function of the main site of production of CSF in humans, the choroid plexus (ChP), *in vitro* and *in vivo*. We captured ChP protein kinetics in human ChP organoids and the CSF of participants with suspected NPH undergoing CSF drainage (n=10) or controls (n=9). We found that transthyretin is abundantly secreted by ChP organoids, and we observe correlations with CSF transthyretin synthesis rates and volume of CSF production *in vivo* (ρ=0.738; p<0.05). Clearance rates of transthyretin are ∼10 fold slower in NPH compared to controls, demonstrating impaired CSF clearance. ChP SILK is a novel clinical tool for interrogating CSF flow.

**One Sentence Summary:** Using stable isotope labeling kinetics of choroid plexus proteins, in human choroid plexus organoids and in vivo, we find that synthesis and clearance of transthyretin is altered in normal pressure hydrocephalus.

## Introduction

Normal pressure hydrocephalus (NPH) is a clinical syndrome characterised by excessive cerebrospinal fluid (CSF) accumulation resulting in ventriculomegaly, gait and cognitive impairment and urinary incontinence (*1*). It is common in the elderly, with an estimated prevalence of around 4% in those over 65 years (*2*).

Little is known about the disease mechanism(s) of NPH, but individuals often respond clinically to CSF diversion through lumbar or ventriculoperitoneal shunting, and a growing body of genetic and functional imaging data supports the notion of it being a distinct disease entity, characterized by abnormal CSF circulation. Two genetic risk factors have been previously identified; SFMBT1 (*3, 4*) and CFAP43 (*5*). Both implicate failure of CSF circulation, but it is not clear whether they impact CSF secretion or reabsorption. A recent GWAS study identified six novel risk genes significantly associated with NPH, several of which have been linked to the function of important fluid barriers in CSF circulation including the blood-brain and blood-CSF barriers (*6*). The only *in vivo* clinical study of CSF flow to date involved injected radio-opaque contrast into the intrathecal space and this demonstrated slower clearance in NPH compared to controls (*7*).

CSF circulation occurs as part of normal brain physiology in humans, with proposed roles in nutrient provision, electrolyte homeostasis and waste clearance. The choroid plexus (ChP) has an important role in CSF production (*8*). It actively secretes and transports water, proteins and other metabolites across the blood-CSF barrier (*9*). It is unclear exactly how CSF and its constituents are cleared from the subarachnoid space, but reabsorption can occur through the arachnoid granulations, and other pathways such as the meningeal lymphatics (*10*) and the glymphatic system (*11, 12*). It is not clear how excessive CSF arises in NPH, but ChP CSF hypersecretion, ependymal denudation, and damage and scarring of intraventricular and parenchymal (glia–lymphatic) CSF pathways have been postulated based on animal studies and observations in humans with acquired hydrocephalus (eg. post-infection or haemorrhage), with a prominent role of neuroinflammation (*11*).

To date, the ability to investigate CSF circulation *in vivo* has been limited by a paucity of appropriate monitoring tools (*11*). Without *in vivo* biomarkers of CSF turnover it has been challenging to: a) accurately diagnose NPH and identify individuals likely to respond to shunt; b) determine whether NPH is associated with dysregulation of CSF production, circulation or clearance; c) develop and interpret static fluid biomarkers of NPH and d) measure transport of proteins across the ChP and blood-CSF barrier, which has important implications for clinical practice and therapeutic intervention. The study of NPH is further confounded by the common co-existence of other neurodegenerative processes with idiopathic NPH (iNPH), notably Alzheimer’s disease (AD) (*13*).

To address these challenges, we set out to develop a novel kinetic protein assay using the stable isotope labeling kinetics (SILK) technique (*14, 15*) to measure the synthesis and clearance rates of ChP relevant targeted proteins in the central nervous system. We hypothesized that proteins abundantly expressed by the ChP could serve as biomarkers of ChP synthetic function or help us understand the disease pathogenesis and potentially identify drug targets. In addition, if these proteins do not easily cross the blood-CSF barrier, they could also serve as biomarkers of CSF circulation and clearance. Conversely, proteins that are not expressed by the ChP or other cells within the brain but are found in abundance in CSF are likely to serve as biomarkers that reflect barrier integrity and their rate of appearance in CSF will reflect their rate of transport across the blood-CSF barrier. In this study, we aimed to characterize and quantify a panel of ChP derived proteins and measure synthesis and turnover rates in human stem cell derived ChP organoids. We then aimed to do the same in humans, compare with organoids and correlate our findings with direct measures of CSF flow *in vivo*.

## Results

### Choroid plexus organoid characteristics

Human choroid plexus (ChP) organoids form fluid-filled sacks that contain clear colourless fluid by day 42 (Fig. 1A). We have previously shown that human *in vitro* ChP organoid fluid (‘iCSF’) has a CSF protein profile similar to human CSF (*16*). The epithelial cells present in the organoid form a polarised barrier, with tight junctions on the apical side and specialised ChP transporters regulating entry of molecules across the barrier. Staining of ChP and CSF transporter markers show that organoids display characteristics of ChP cells by day 40 *in vitro* (Fig. 1B). Previously published single-cell RNA sequencing of ChP organoids show development and maturation of distinct cell populations including ChP epithelial and stromal cells, as well as some neuronal progenitors and neurons (*16*). Single-cell RNA sequencing between day 27 and day 53 showed enrichment of ChP epithelial and stromal cells from the ChP in ChP organoids compared to control, unguided cerebral organoids (Fig. 1E-F). We also show expression of albumin, cystatin-C (Cys-C) and transthyretin (TTHY), indicating abundant expression of Cys-C and TTHY and very little expression of albumin (Fig. 1G).

**Fig. 1.**
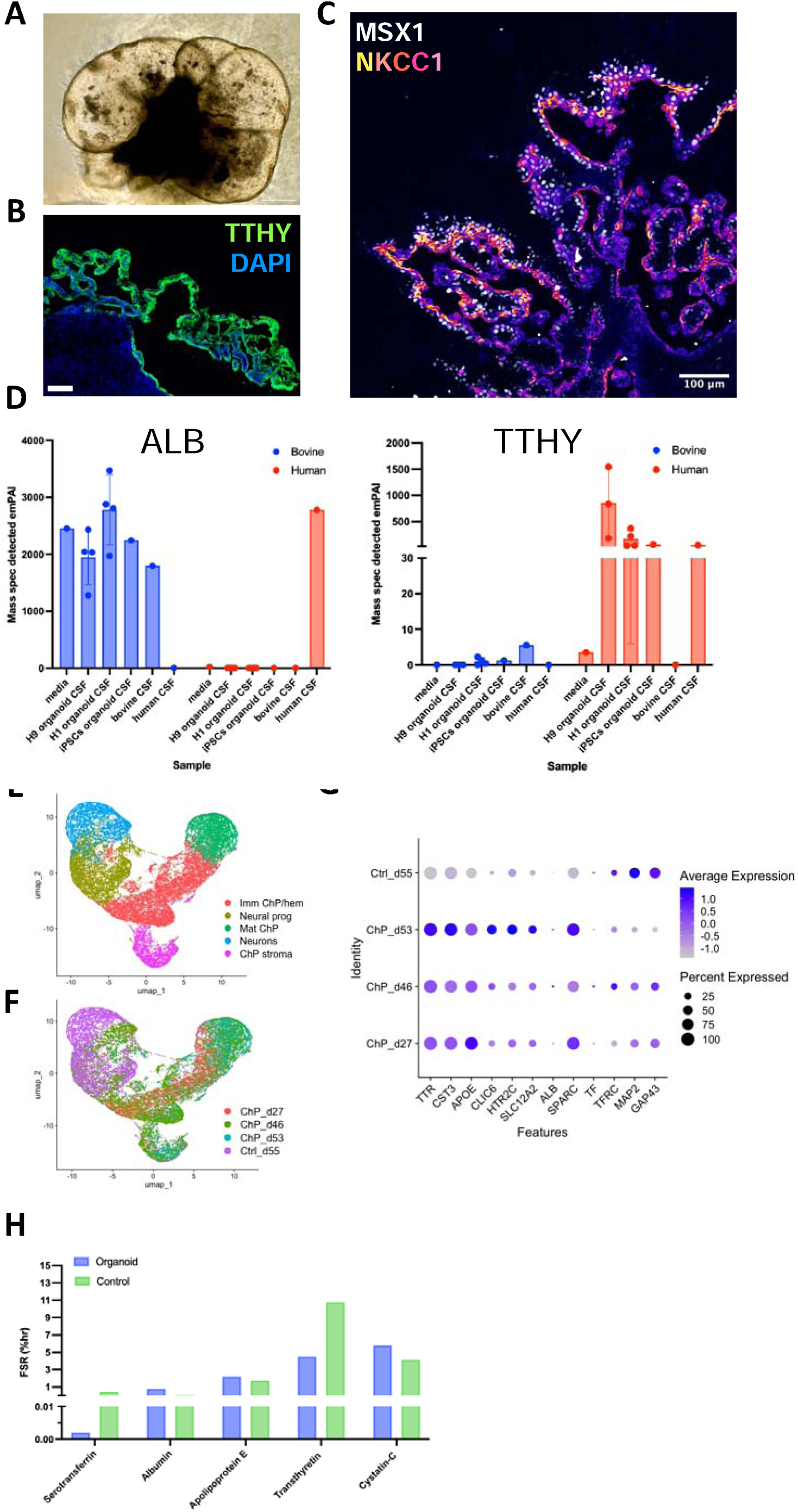
Choroid plexus organoid characteristics. (**A**) Bright field image of a H1 ChP organoid at day 42 that developed self-contained fluid compartments with CSF-like fluid. Scale bar, 250μm. (**B**) Representative confocal image of whole ChP organoid from H1 at day 40 stained for TTHY in green and DAPI in blue. Scale bar, 100μm. (**C**) Representative confocal images of ChP epithelium from a H9 derived organoid at d60 stained for ChP marker MSX1 and CSF transporter NKCC1, nuclei are stained with DAPI (blue). Scale bar, 100μm. (**D**) Bar charts with scatter plots of mass spectrometry detected emPAI values for ALB and TTHY peptides of human and bovine origin from organoid media, organoid CSF from 3 independent cell lines (human embryonic lines H9, H1 and iPSC line IMR90-4), bovine foetal CSF and human adult CSF (mass spectrometry data from Pellegrini et al.(*16*)). (**E**) Single Cell RNA (scRNA) sequencing of ChP organoids (ChP day27, ChP day46, ChP day53) and cortical organoid (Control day55) showing the different cell populations: ChP stroma, neurons, ChP epithelial cells (Mat ChP), hem/ChP progenitors (Imm ChP/hem) and neuronal progenitors (Neural prog). (**F**) scRNA sequencing showing different timepoints analysed (ChP day27, ChP day46, ChP day53, Ctrl day55). (**G**) Dot plot showing average expression and percentage of cells expressing the displayed enriched genes identified by scRNA sequencing (scRNA seq data from Pellegrini et al.(*16*)). (**H**) Bar chart comparing FSR% (log^10^ transformed) choroid plexus proteins synthesized by organoid (blue) labeled by SILK, and human controls labeled by SILK (green). ALB, albumin; ChP, choroid plexus; CSF, cerebrospinal fluid; FSR, fractional synthesis rate; NKCC1, Na+/K+/2Cl− cotransporter; NPH, normal pressure hydrocephalus; TTHY, transthyretin.

We selected a panel of proteins that were highly abundant in both iCSF and human control CSF, known to be translated by, or transported across ChP epithelial cells, and that were also amenable to SILK (contain leucine residues). The proteins selected were albumin, apolipoprotein E (ApoE), Cys-C, serotransferrin and TTHY. Since some of these proteins are also constituents of organoid development media, we used differences in sequence homology with targeted proteomics to differentiate proteins of human origin from bovine (Fig. 1D). Albumin peptides in iCSF consisted mostly of bovine albumin, indicating that albumin was transported from the media across the media/CSF barrier. TTHY in iCSF was almost exclusively human indicating that it was translated by organoid cells.

### Human subject characteristics

Individuals were recruited to measure protein kinetics *in vivo* using SILK. Subject characteristics are summarised in Table S1. A total of nineteen individuals were included in this study; ten individuals with suspected NPH, four individuals recovered from the acute phase of subarachnoid haemorrhage (‘SAH-controls’) where ventricular CSF was collected (Control [V]); and five controls from a validation control cohort, where lumbar CSF was collected. This cohort was composed of five amyloid negative age-matched controls (Control [L]).

Comparing the NPH and SAH-control groups, the controls were younger (NPH: 75, (71-78) vs 57 (46-65) years old; *p* = 0.0012). There was no significant age difference between NPH and lumbar control cohorts (NPH: 75 (71-78) vs 70 (63-84) years old; *p*=0.2438). The sex distribution of the NPH and SAH-control groups also differed between groups. The entire NPH cohort had abnormal gait on examination and the majority of individuals reported urinary incontinence (80%) (Table S1). Most individuals with NPH reported cognitive symptoms; 70% reported subjective impairment of episodic memory and 50% problems with executive function. Objective cognitive screening showed cognitive impairment to be mild (MMSE: median 28; IQR 26-28; n=9). None had CSF biomarker support for amyloidosis. All individuals had imaging features of disproportionately enlarged subarachnoid-space hydrocephalus, determined by a clinical neuroradiologist. 5/10 had CT imaging before and after shunting, and demonstrated reversibility of ventriculomegaly following shunt insertion with a mean change in ventricular volume of −27.5 ml; −13.11% change (Fig. S1). Seven suspected NPH patients responded to CSF diversion with an improvement in gait and/or cognition. Two did not respond and one participant died of other unrelated causes and the surgical outcome could not be determined.

### ChP protein kinetics in human choroid plexus organoids

Using SILK proteomics and plotting the tracer-to-tracee ratio (TTR) of peptides corresponding to the ChP-related proteins of interest at several timepoints during- and post-labeling (Fig. S2A), we captured the turnover of all five proteins in ChP iCSF including plasma derived proteins albumin and serotransferrin (Fig. S2B-C and G-H); and ChP derived proteins TTHY, Cys-C and ApoE (Fig. S2D-F and J-K).

The rate of labeling of plasma derived proteins in iCSF was very low, with an albumin TTR of ∼1.5% (FSR 0.004 %/hr), and serotransferrin TTR of <0.4% (FSR −0.01 %/hr) indicating very low synthesis and/or low release into iCSF. In organoid lysate, the TTR of albumin was ∼10% (FSR 0.8 %/hr), suggesting some albumin is synthesized by ChP organoid cells. For serotransferrin the TTR was 0.15% (FSR 0.002 %/hr), indicating negligible synthesis by ChP cells.

By contrast, the TTR of peptides corresponding to ChP derived proteins is around 20-50 fold higher, depending on the protein (Fig. 2C-G): TTHY (iCSF peak-of-labeling TTR ∼50%; FSR 4.51 %/hr; organoid lysate peak TTR 50%; FSR 4.0%/hr), Cys-C (iCSF peak TTR ∼60%; FSR 5.78 %/hr; organoid lysate peak TTR ∼50%; FSR 2.8 %/hr) and ApoE (iCSF peak TTR 35%; FSR 2.20 %/hr; organoid lysate TTR 20%; FSR 1.2 %/hr).

**Fig. 2.**
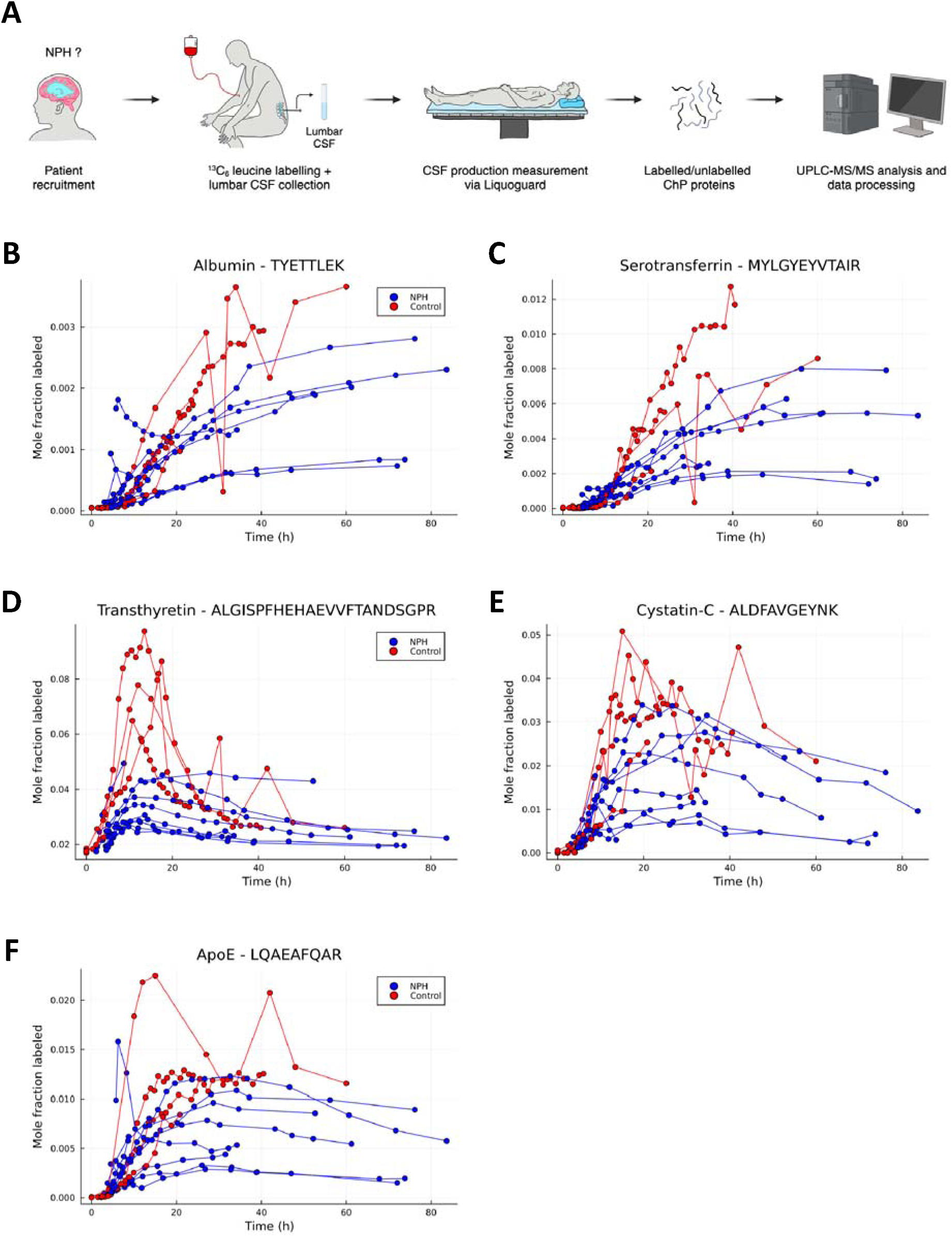
Choroid plexus protein turnover in human CSF: plasma and ChP derived proteins. (**A**) Schematic of SILK labeling method *in vivo*. (**B-F**) *In vivo* SILK time course profiles of ChP proteins in both NPH and control CSF for (**B**) Albumin (TYE peptide) (**C**) Serotransferrin (**D**) TTHY (ALG peptide) (**E**) Cys-C (**F**) ApoE. ApoE, Apolipoprotein E; ChP, choroid plexus; CSF, cerebrospinal fluid; Cys-C; Cystatin-C; NPH, normal pressure hydrocephalus; SILK, stable isotope labeling kinetics; TTHY, transthyretin. Fig. 2A was generated using BioRender.com.

As expected, the FCR of all proteins is extremely low (<0.1 %/hr) in iCSF and lysate, indicating that no significant protein clearance occurs in either compartment.

### ChP protein kinetics *in vivo*

In the time window studied (72 hours), we captured the fractional synthesis (FSR) and fractional clearance (FCR) rates of TTHY, Cys-C and ApoE. TTHY had the highest FSR, followed by Cys-C and ApoE (summarised in Table S2). Serotransferrin and albumin had much lower FSR estimates that continued to rise at 72 hours – thus, only synthesis rates could be measured for these two proteins. FSR was significantly higher in controls compared to NPH for both the ALG and TSE peptide monitored for TTHY (p<0.05) and serotransferrin (p<0.05).

FCR was captured for TTHY, Cys-C and ApoE. For TTHY, FCR was ∼10-fold lower in NPH than controls (ALG peptide: p<0.01; TSE peptide: p<0.01). For comparison, the kinetic curves of all five proteins within single NPH and control subjects are shown in Fig. 3A-B.

**Fig. 3.**
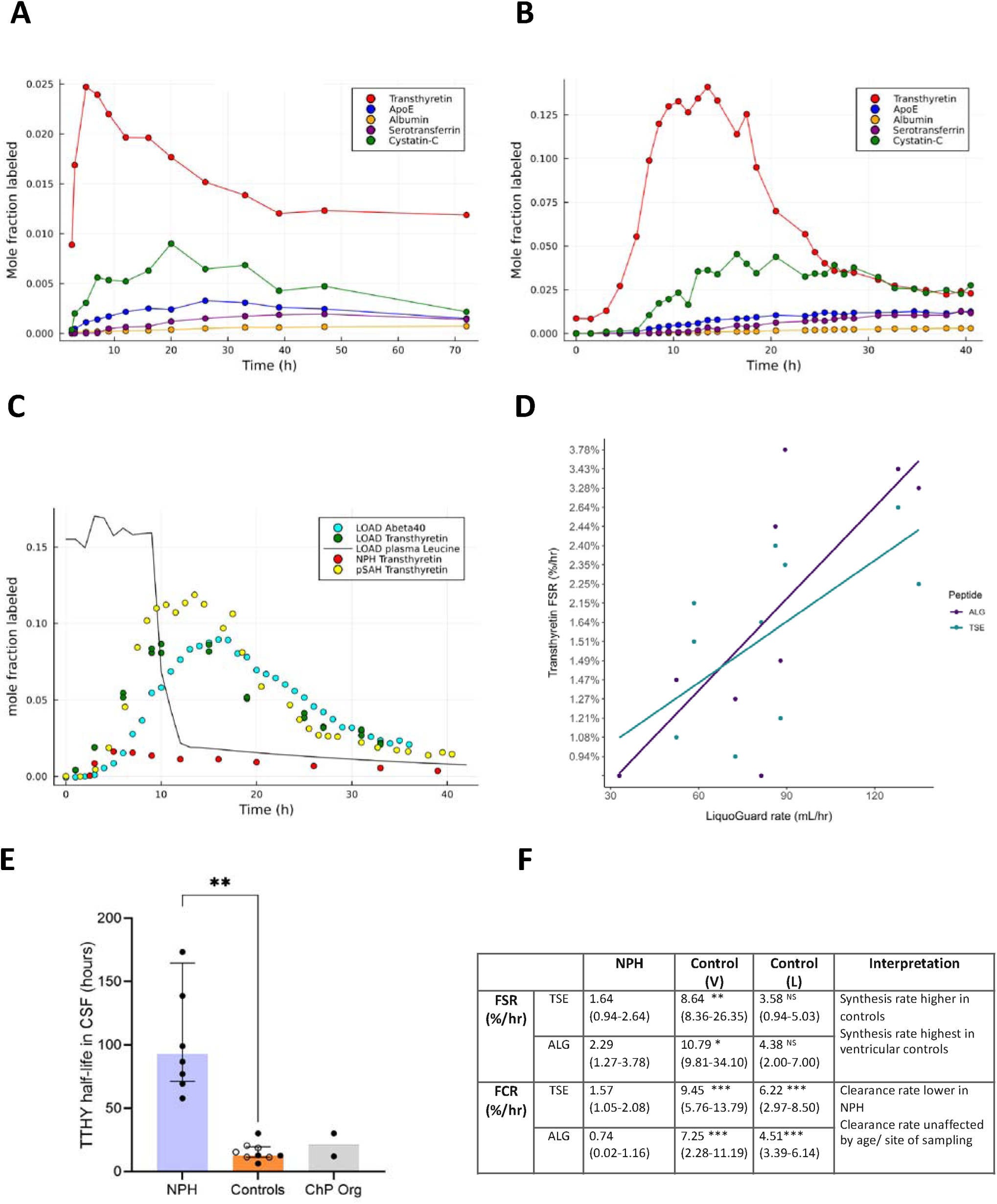
Summary of ChP kinetics in vivo. (**A**) Kinetic curve of five peptides within a single subject (NPH). (**B**) Kinetic curve of five peptides within a single subject (control). (**C**) Comparison of ChP kinetics in ventricular and lumbar CSF. (**D**) Correlation of fractional synthesis rate of TTHY peptides ALG (p=0.037, ρ=0.738, R^2^=0.483) and TSE (p=0.042, ρ=0.683, R^2^=0.519) with CSF production rate by LiquoGuard. (**E**) TTHY turnover in NPH patients (n=8), Controls (Control [V] and Control [L]) (n=9) and ChP organoids (n=2). Bars represent the median +/− interquartile range. Control (L) are depicted with circles. Control (V) are depicted with solid black circles. ** Represents significance at p<0.0021 (unpaired t-test with Welch’s correction). (**F**) TTHY TSE peptide fractional synthesis rate and fractional clearance rate summary *in vivo;* median (range). Significance between NPH and each control group is represented as follows: * p<0.05; ** p<0.01; ***p<0.005; NS = Not significant. FSR, fractional synthesis rate; FCR, fractional clearance rate; L, lumbar; V, ventricular.

### Comparing ChP kinetics in ventricular and lumbar CSF

Since SAH-control CSF was ventricular and NPH CSF was lumbar, we investigated whether the site of CSF collection could confound SILK measurements. We compared FSR and FCR values for TTHY between the study’s subarachnoid haemorrhage (SAH) controls (yellow-*ventricular*) and a validation control group ‘lumbar controls’, who were amyloid negative age-matched controls who donated lumbar CSF (dark green-*lumbar*). The results showed strong alignment of the kinetic curves derived from these two control groups, despite their CSF compartmental differences (a single subject shown in Fig. 3C; other individuals shown in Fig. S4). This makes it unlikely that differences found in controls *vs* NPH could be due to differences in kinetics between the two CSF compartments. Although the analytical approaches at each SILK site utilised mass spectrometry, direct comparison between the NPH and validation controls was not performed due to slight method variations across sites. However, an exploratory analysis comparing TTHY FCR between the two groups showed a lower TTHY FCR in NPH compared to controls in lumbar CSF (Table S2). We also note that TTHY FSR was higher in ventricular controls compared to lumbar controls.

### Relating protein kinetics to measured CSF flow

In NPH subjects, the FSR of TTHY for both peptides monitored was found to positively and significantly correlate with CSF production as measured by LiquoGuard7 in-clinic (p=0.037, ρ =0.738 for ALG peptide and p=0.042, ρ=0.683 for TSE peptide) (Fig. 3D). No association was observed between the rate of CSF production by LiquoGuard7 and the FSRs of Cys-C, serotransferrin, albumin and ApoE (p values = 0.5518, 0.8737, 0.8024 and 0.4194, respectively). This suggests that the FSR of TTHY in CSF could be a biomarker reflective of CSF production.

## Discussion

We introduce a novel SILK method to measure the synthesis and clearance of proteins in human CSF, revealing significant impairment in CSF clearance in NPH compared to controls. We also observed disruption of choroid plexus (ChP) synthetic function, suggesting a possible feedback loop that may regulate CSF synthesis in humans.

Since there is no pathological hallmark of NPH (*17*) and still doubt that NPH is a distinct disease entity (*18*), we set out to determine if we could detect objective differences in protein kinetics in this population. We designed a panel of novel kinetic protein targets that would allow us to track a) the synthesis of proteins within ChP epithelial cells into CSF; b) the transport of peripherally-derived proteins across the blood-CSF barrier and c) the clearance/reabsorption of proteins from CSF. There are established SILK methods for quantifying synthesis and clearance rates of Aβ, tau, APP (*14, 15*) and here we report an adapted method to determine the kinetics of proteins derived from, and reflective of the ChP function *in vitro* and *in vivo*. We found that the synthesis rate of TTHY, or the retinol or thyroxine it carries, correlated with a direct measure of volume of CSF synthesized, suggesting this method could be an *in vivo* biomarker of CSF flow.

We used human ChP organoids to determine which proteins were most abundantly expressed by ChP epithelial cells. Based on previous work using untargeted mass spectrometry of organoid CSF and human CSF (*16*), paired with single-cell RNA sequencing to confirm which cells they derived from, we were able to establish that TTHY, Cys-C and ApoE were abundantly expressed and rapidly translated by ChP epithelium. Importantly, we found similar results in human CSF. We found extremely low turnover rates *in vivo* and *in vitro* for albumin and serotransferrin, indicating that they are likely to be produced peripherally by the liver and then transported across the blood-CSF barrier.

This isotope labeling method offers a novel tool to quantitate the synthetic function of the ChP *in vivo* and its capacity to transport proteins or drugs across the blood-CSF barrier. Since these findings are recapitulated in ChP organoids, with broadly similar synthesis (FSR) rates, this model has potential utility for understanding ChP physiology in a dish, and interrogate diseases of abnormal ChP physiology such as ChP tumours and idiopathic intracranial hypertension. ChP organoids may also have wider utility in testing factors which influence therapeutic drug delivery to the central nervous system.

We also aimed to develop a method capable of quantitating CSF protein clearance or resorption. Of the proteins identified, TTHY was of particular interest as it is produced mainly by the ChP (*19*), is rapidly translated and does not readily cross the blood-CSF barrier, making it an attractive SILK target to monitor CSF clearance. Other proteins like ApoE are known to be expressed by other cell types within the CNS (*20*). TTHY is a protein with a molecular mass of 55kDa with a tetrameric structure. Physiologically it is involved in transporting the thyroid hormone thyroxine into the brain along with retinol binding hormone (*21*). The two primary sites of TTHY synthesis are the liver (*22*) and the ChP (*23*) which give rise to TTHY in the plasma and CSF, respectively. However, there is an 11-fold difference in the mRNA levels and a 13-fold difference in the speed of synthesis between the two regions with faster production occurring in the ChP (*24*). Since rapid labeled leucine incorporation is seen both in organoids (where there is no other TTHY source) and *in vivo*, we conclude that most TTHY expression in CSF is derived from ChP. We next considered whether TTHY clearance was likely to be a good marker of CSF bulk flow clearance or whether it had other routes for removal. Unlike amyloid beta (Aβ), which may be cleared via the coordinated action of ApoE and the low-density lipoprotein receptor-related protein 1 (LRP1) (*25–27*) and possibly other transporters such as p-glycoprotein (PGP) (*28*), removal of TTHY does not appear to be aided by transport proteins. To our knowledge, mechanisms of TTHY clearance from the brain have not been well studied. A limited number of studies have assessed the ability of TTHY to cross the blood-brain barrier (BBB) but have done so in the context of either TTHY-assisted Aβ transport (*29*) or TTHY-iododiflunisal (IDIF) transport (*30*). Both studies suggested that TTHY has the ability to cross the BBB but this likely only occurs in the brain-to-blood direction (*29*).

The most striking difference between NPH and control subjects is the FCR (rate of clearance) of TTHY, which is ∼10 times lower in NPH. This was validated by monitoring a second TTHY peptide and in two independent control groups. Although it did not reach significance, a similar trend was seen for Cys-C. We considered whether it could be an artefact of the site of CSF collection (controls had ventricular CSF sampling), and validated the results on a further independent control group of five cognitively normal, amyloid negative, age-matched individuals from another institution (Washington University, St Louis) for whom isotopically labeled lumbar CSF was available. Since this cohort were labeled using the same clinical protocol, were age-matched, and had CSF collected in the same way and from the same site, they represented an ‘ideal’ validation opportunity. This indicates that CSF protein turnover is ∼10 fold longer in NPH, suggesting possible fluid and protein stasis and greater potential for protein aggregation and/or post-translational modification.

Failed CSF clearance has become an important theme in NPH research, but the mechanism by which failed clearance of CSF occurs is yet to be established and there are currently no clinically available tools to capture this, other than imaging intrathecal contrast clearance, which is significantly delayed in NPH (*7*). Pathways of waste removal from the CNS has been a subject of investigation in recent years (*31*). The conventional view of brain fluid clearance and waste removal supports clearance via three routes; via arachnoid granulations into the dural venous sinuses; via nasal lymphatics and the cribriform plate into the cervical lymph nodes (CLNs) and via transporters/receptors located in the ChP epithelium (*32*). In contrast to other tissues, the parenchyma of the brain lacks a typical lymphatic vascular network (*10*). Importantly, several studies have recently investigated and characterised the meningeal lymphatic vessels (*10, 33, 34*). Removal of cellular waste products, which in the periphery is normally cleared by the traditional lymphatic network, is partly fulfilled by CSF-ISF exchange paravascular (glymphatic) route (*10, 12, 32*). The glymphatic route, first described in 2012 (*12*), is comprised of a network of perivascular channels which drain into the meningeal lymphatic vasculature or the dural sinuses (*35*). MRI studies have generated evidence to support that glymphatic clearance of brain fluid is impaired in individuals with NPH (*7, 36*). This may be perpetuated by the loss of neuronally-derived ionic waves in brain interstitial fluid (*37*). Interestingly, in one such study, parenchymal glymphatic enhancement peaked overnight and this effect was attributed to heightened glymphatic function during sleep(*36*). Furthermore, reduced tracer clearance from the entorhinal cortex has been observed in an NPH cohort (*38*). It has been suggested that the combined dysfunction of the glymphatic and meningeal systems may promote ventricular reflux (*39*). We speculate that impaired glymphatic and meningeal function may result in reduced ChP protein/TTHY clearance.

We do not yet know which biological process(es) lead to CSF protein clearance impairment in NPH, and this should be the direction of future basic science studies. However, hydrocephalus can be induced in rodent models that receive intraventricular autologous blood, and hydrocephalus can be subsequently attenuated by complement inhibitors (*40*). In patients, insults known to induce the classical complement cascade are common clinical causes of secondary hydrocephalus; post-infectious and post-haemorrhagic hydrocephalus are the leading causes of childhood hydrocephalus worldwide (*41*).

We also observed significant differences in ChP protein translation in NPH compared to controls. The FSR of TTHY and serotransferrin were significantly lower in NPH providing *in vivo* evidence of reduced epithelial cell protein translation (TTHY) and reduced active transport across the ChP (serotransferrin). This may indicate that a feedback mechanism exists to regulate the ChP in humans.

Finally, we found a relationship between TTHY FSR and the measured volume of CSF production. We recognize that the movement of water and protein in the CSF may be partially independent of one another. Around 50% of water in the CSF compartment passes into this space by osmosis, and this process is driven by the osmotic draw of CSF proteins. Water can also be transported into the CSF space against an osmotic gradient (*42*) through active transport notably by the Na+/K+/2Cl− cotransporter (NKCC1) expressed in the luminal membrane of ChP cells (*43*). Since it is practically difficult to label and track water *in vivo* we chose to correlate TTHY, a protein marker of ChP synthetic function, with a measure of CSF production. The significant correlation between them suggested that ChP SILK may be a valuable surrogate marker for tracking CSF flow *in vivo*. The significant correlation between TTHY and the volume of CSF production highlights the potential of this method as a surrogate marker for tracking CSF dynamics in NPH and other conditions where CSF flow is disrupted.

Labeling ChP proteins using SILK could be a useful clinical diagnostic marker of NPH. Prolonged diagnostic lumbar drainage is currently the gold standard diagnostic test to determine which individuals are likely to clinically respond to CSF diversion (*44*) and requires a three-day admission to hospital making it costly and burdensome for patients and healthcare providers. Labeling patients with a stable isotope prior to large volume CSF tap could provide objective evidence of clearance failure in addition to clinical evidence of gait improvement to provide a more accurate predictive test of clinical responders who might benefit from ventriculoperitoneal shunting, at a lower cost. This study was not designed to use ChP SILK to assess the clinical value of SILK to determine shunt response, and the majority (7/9) of individuals improved in response to CSF diversion, meaning we lacked a negative control group.

There is likely to be considerable interest in a kinetic protein biomarker that reflects CSF flow in other areas of neuroscience. Notably, in intrathecal therapeutics where compounds such as antisense oligonucleotides are administered directly into CSF given that the rate at which an individual circulates CSF is likely to influence drug delivery, clinical efficacy and drug safety. A number of ASO studies have been terminated early owing to safety concerns. The Htt ASO trialled in Huntington’s disease was terminated in part due to ventriculomegaly (*45*). This could have been a manifestation of accelerated neurodegeneration with accelerated volume loss. The other explanation is that they developed stasis of CSF circulation caused by increased CSF protein concentration or inflammation.

This study has some limitations. A major challenge in studying NPH is the lack of diagnostic biomarkers. We selected cases that had clinical features that met international criteria for probable NPH (*46*), and radiological support for disproportionately enlarged subarachnoid-space hydrocephalus for NPH. All individuals were examined by a board-certified neurologist prior to inclusion to screen for other more likely diagnoses (eg. progressive supranuclear palsy). We were also able to establish that 5/5 cases tested had reversibility of ventricular volume following ventriculoperitoneal shunting, which is strongly associated with clinical reversibility (*47*). Other limitations include small population size (n=19) and that cases and controls were recruited from different clinical centres. SILK studies are expensive and labour intensive and so this still represents the largest study of this type in NPH. Average age of the two groups was significantly different and the control cohort were younger than the NPH group. Previous studies have demonstrated that Aβ kinetics decline with age (*48*). We cannot exclude the possibility that ChP protein clearance may also slow with increasing age. Access to serial labeled control CSF is challenging and so the control group, individuals who had another disease process – acute subarachnoid haemorrhage, could potentially have influenced differences in the results. We were able to correct for this potential confounder by identifying a second age matched control group. These samples were also derived from ventricular CSF rather than from the lumbar space. The second validation control group had lumbar CSF samples labeled at another site by SILK and these results verified that SILK curves closely resembled those of the other controls. Both control cohorts had been analysed using the same mass spectrometer and analytical pipeline and so they could be compared directly. We were interested to note that the synthesis rate of TTHY between control groups was different, with higher synthetic rates in the younger SAH ventricular controls; this aligns with suggestions that the synthetic capacity of the ChP may decline with age (*49*).

In conclusion, our SILK method provides a valuable tool for quantifying ChP function and CSF protein dynamics *in vivo* and *in vitro*. ChP organoids recapitulate human ChP cells and highlight the utility of this model in studying diseases of CSF disruption and evaluating intrathecal drug delivery. The identification of TTHY as a potential kinetic biomarker of CSF flow *in vivo* offers a novel approach for diagnosing and monitoring NPH. This is likely to be a valuable clinical tool for quantitating CSF flow in the diagnosis of NPH and for detecting other disruptions to CSF flow due to intrathecal drug delivery. We also see utility in this method for investigating impaired CSF flow as a priming event for the formation of other age associated proteinopathies, such as AD. Future research should explore the broader applicability of these biomarkers in neurodegenerative conditions and their potential role in predicting response to therapeutic interventions.

## Materials and Methods

### Stable Isotope Labeling Kinetics (SILK) in ChP organoids

#### Generation of ChP organoids

Human H1 ES cells were obtained from WiCell and used for the generation of choroid plexus organoids using Stem Cell Technologies Cerebral Organoid kit (catalog nr. 08570 and 08571) reagents, as previously described (*16*). Briefly, EBs were generated by seeding 4000 cells in a 96-well U bottom low attachment plate with EB media and 50 μM Y-27632 ROCK inhibitor for 3 days. On day 5, the culture media was switched to NI (neural induction) within the same 96-well plate. On day 7, EBs were embedded in 30 μl of Matrigel (Corning) using dimpled parafilm sheets, following the established procedure described by Lancaster et al. (*50*), and incubated for 20min at 37°C. Subsequently, the EBs were transferred to a 6-well plate, with each well containing 3ml of Expansion media. For ChP patterning, 3 μM CHIR and 20 ng/ml BMP4 were added in Maturation media on day 10 until day 17. Starting from day 30, dissolved Matrigel (at a ratio of 1:50) was introduced to the Maturation media.

#### ChP profiling by single cell RNA sequencing

Organoid single-cell dissociation, library preparation and sequencing were all previously described in Pellegrini et al. (*16*). Briefly, single-cell dissociation was performed by first pooling two organoids for each condition: 55-day H9 telencephalic organoids, 27-day H1 ChP (ChP sample 1), 46-day H1 ChP (ChP sample 2), and 53-day H1 ChP (ChP sample 3) into a 15 mL tube. Samples were incubated in 1ml of Accumax (Sigma, A7089) with 400 μg DNase I and 15 μM actinomycin D at 37°C for 20min with gentle agitation. At 5min intervals, the sample tubes were flicked then pipetted 10 times. Clumps were allowed to settle and supernatant collected, to which 100 μL FBS was added before filtration on a 35μm filter tube (Corning, 352235). Samples were then spun at 300*g* for 5min. Dead Cell Removal kit and MACS column (Miltenyi, 130-090-101) were used to remove dead cells before another spin at 300*g* for 5min. Cells were resuspended in an appropriate volume of 0.04% BSA in PBS to load 16,000 cells per well on the 10X Chromium system (10X Genomics). Raw and processed human organoid sequencing data are available at the Gene Expression Omnibus (GEO) database GSE150903.

#### ChP organoid SILK

ChP organoids were labeled with 50% mol (100% TTR) ^13^C_6_-leucine for one week. Following labeling, organoids were washed with fresh, unlabeled media and subsequently incubated with regular maturation media for a 24hr chase (Fig. S2A). CSF-like organoid fluid (iCSF) was collected at multiple time points (1hr, 6hr and 24hr). For collecting iCSF, organoid media was removed and organoids washed with fresh media. iCSF was then extracted using a pulled glass microcapillary as previously described (*16*). Briefly, iCSF was collected using a pulled glass microcapillary attached to filter and tubing using controlled suction. iCSF was centrifuged at 10,000*g* for 10min to pellet debris before supernatant was collected, snap frozen in liquid nitrogen and stored at −80°C. iCSF samples were prepared for mass spectrometry analysis using the same protocol as CSF described below.

### ChP Stable Isotope Labeling Kinetics (SILK) in human subjects

#### Recruitment and SILK protocol for NPH participants (NPH cohort)

The NHS Health Research Authority, Research Ethics Committee London-Bloomsbury gave ethical approval for the UCL NPH SILK study. Individuals with suspected idiopathic NPH (iNPH) were recruited from the specialist hydrocephalus service at the National Hospital for Neurology and Neurosurgery, Queen Square, London. Informed written consent was given. Individuals were prospectively assessed and examined by a board-certified neurologist, and specific symptoms and examination features were recorded according to a pre-specified questionnaire. A collateral history was obtained from their study partner where possible. A timed walk was carried out by a member of their clinical team.

Individuals then underwent SILK as previously described (*14*). Briefly, participants were labeled intravenously with ^13^C_6_-leucine at a concentration of 8.5 mg/mL prior to undergoing diagnostic lumbar drainage, with leucine administered at a rate of 3 mg/kg/hr for 10 minutes, followed by 2 mg/kg/hr for up to 9 hrs. The infusion was continued until drain insertion, thus the overall duration of infusion varied dependent on clinical operational factors which influenced the time the individual was taken to surgery.

To correct for variation in infusion duration and other physiological factors that might influence leucine metabolism, tracer-to-tracee ratios were adjusted for infusion duration using plasma leucine enrichment at plateau, measured using HILIC-MS/MS (detailed below). Research CSF was removed regularly for the duration of the diagnostic drainage period. CSF was collected in a 10 mL syringe and transferred to a polypropylene Falcon container and centrifuged at 4°C for 10 minutes at 1,500*g*. Samples were then aliquoted into 1.5 mL microcentrifuge tubes and stored at −80°C prior to UPLC-MS/MS analysis.

#### Recruitment and SILK protocol for control participants (SAH cohort; ‘Control (V)’)

Controls were patients with non-traumatic subarachnoid hemorrhage at the Centre Hospital Montpellier, who had recovered from their acute illness and CSF flow rate had plateaued to within normal clinical parameters as determined by their treating clinicians. Their ventricular shunts remained in-site for an additional 72 hours on a research basis to facilitate SILK. Leucine was administered at a rate of 3 mg/kg/hr for 10 minutes, followed by 2 mg/kg/hr for 9 hrs. CSF was withdrawn at regular intervals and processed prior to storage at −80°C as described above.

#### Recruitment and SILK protocol for lumbar control participants (Amyloid negative cohort; ‘Control (L)’)),

Controls were research volunteers, who had a research lumbar drain placed at Washington University St Louis, as previously described (*14, 51*). Leucine was administered at a rate of 3 mg/kg/hr for 10 minutes, followed by 2 mg/kg/hr for 9 hrs. CSF was withdrawn at regular intervals and processed prior to storage at −80°C as described above.

#### Clinical measurement of CSF flow in NPH subjects

Lumbar drain (LD; Medtronic® Duet epidural catheter, USA) was inserted in-between the L3/L4 spinal lumbar vertebrae. LiquoGuard7 (MÖLLER Medical GmbH, Germany) was attached to the drainage catheters and the LiquoGuard7 external drainage tubing was primed using sterile normal saline to prevent CSF wastage.

The external intracranial pressure (ICP) transducer of the LiquoGuard7 pump was applied on the body of the patients in line with the external auditory meatus. Participants were requested to remain lying flat for the total duration of the procedure, which included an initial 20 mins for CSF flow rates to stabilise and an additional 30 mins for the calculation of the CSF production rate. The LiquoGuard7 parameters were set to bring the ICP pressure to 0mmHg and to allow CSF to be drained freely into the external drainage bag of the LiquoGuard7 pump at this low ICP. The LiquoGuard7 software was used to analyse the flow rate data and calculate an hourly CSF production rate (mL/hr).

#### Neuroimaging analysis of NPH subjects

Participants had brain imaging (either magnetic resonance imaging or computerised tomography (CT) as part of their normal clinical care, either to confirm diagnosis or for monitoring for complications. Using a deep-learning-driven CT-brain quantification pipeline (*52*), 10 pre- and post-shunt CT scans from 5 unique participants underwent brain segmentation analysis. This pipeline enables a fully automated quantitative segmentation of brain structures, including VCSF and ICV in brain CT images. Employing trained deep learning models, the pipeline efficiently segments various tissue classes within the input CT images. Subsequently, segmented VCSF and ICV maps are binarized, and their volumes are extracted. From these volumes, an iNPH-related CT-based volumetric measure, VCSF/ICV, analogous to the 3D version of Evans’ Index, was derived. Further, the changes in CT-VCSF volume and CT-VCSF/ICV are visualised pre- and post-shunt, and over days after shunt.

#### Measurement of plasma ^13^C_6_-leucine enrichment by HILIC-MS/MS

To quantitate labeled leucine enrichment in plasma, a hydrophilic interaction liquid chromatography – tandem mass spectrometry (HILIC-MS/MS) assay was adapted from Prinsen et al. (*53*) to measure ^13^C_6_-/^12^C_6_-leucine ratios, with the following optimisations and adaptations.

Analysis was performed using an Acquity H-Class Ultra Performance Liquid Chromatography (UPLC) system, fitted with an ACQUITY UPLC BEH Amide column (100A; 1.7µm, 2.1 x 50mm) attached to a VanGuard UPLC BEH Amide precolumn (2.1 x 5 mm), which was coupled to a Xevo TQ-S triple quadrupole mass spectrometer operated in positive electrospray ionisation (ESI^+^) mode (Waters, UK). Chromatographic separation was performed over a 10-minute HILIC gradient using mobile phases A (10 mM ammonium formate in 85% ACN 0.15% FA) and B (10mM ammonium formate in ultrapure Milli-Q water 0.15% FA). The column was primed and equilibrated for 45 minutes in initial conditions (100% A at 0.4 mL/min) and kept at 35°C. At injection (1 μL/sample), the column was kept in initial conditions for 3 minutes until a linear gradient of increasing %B began. From 3 – 3.1 minutes B was increased to 5.9%, and from 3.1 – 5 minutes was increased to 17.6%, followed by a final increase to 29.4% from 5 – 6 minutes. The flow rate was then increased to 0.6 mL/min and the column re-equilibrated in 100% A for 4 minutes. Mass spectrometer parameters were set as follows: source temperature (150°C), capillary voltage (1.00 kV), desolvation temperature (550°C), desolvation gas flow (1000 L/hr) and cone gas flow (150 L/hr).

Ion transitions for ^12^C_6_-leucine (*precursor: 132.102 m/z, product: 86.100 m/z*), ^13^C_6_-leucine (*precursor: 138.122 m/z, product: 91.139 m/z*) and ^13^C_6_-,^15^N_2_-lysine internal standard (*precursor: 155.127 m/z, product: 90.100 m/z*) were analysed by multiple reaction monitoring (MRM) and acquired data imported into Skyline (MacCoss Lab, Seattle, USA) for processing and peak integration. Peak areas were exported into Microsoft Excel and leucine TTR calculated as molar ^13^C_6_-leucine/^12^C_6_-leucine peak area ratios.

#### UPLC-MS/MS analysis of ChP peptides in iCSF and CSF

NPH and control SAH CSF samples were thawed on ice before 350 µL of CSF was spiked with 150 ng of yeast enolase internal standard (E6126, Sigma-Aldrich). Protein was precipitated by addition of 3-sample volumes of ice-cold acetone, which was incubated at −20°C overnight (16 hrs). Samples were centrifuged at 16,000*g* for 20 minutes at 4°C, the acetone supernatant removed and the protein pellet allowed to air dry. The pellet was then resolubilised in digest buffer (6 M urea, 2 M thiourea, 2% ASB-14, 200 mM Tris-HCl, pH 8) for at least 20 minutes before reduction and subsequent alkylation of peptides by incubation with 1,4-dithioerythritol (DTT; Sigma-Aldrich, UK) and iodoacetamide (IAA; Sigma-Aldrich, UK) respectively. Two µg of Trypsin-LysC (MS grade, Promega) was added to each sample followed by incubation at 37°C overnight (16hrs). Tryptic peptides were purified by C18 solid-phase extraction (Bond Elute, Agilent Technologies, UK). Purified peptides were lyophilised using a centrifugal evaporator (SpeedVac, Eppendorf) and reconstituted in 50 µL 3% acetonitrile (ACN) 0.1% formic acid (FA) prior to UPLC-MS/MS analysis.

Targeted analysis of the ChP protein SILK panel *in vitro* and *in vivo* was performed by tandem mass spectrometry on an Acquity I-Class PLUS UPLC system coupled to a Xevo TQ-XS mass spectrometer operated in positive electrospray ionisation (ESI+) mode (all Waters, UK). A multiplexed assay was developed and optimised to monitor peptides that were proteotypic for human transthyretin, cystatin-c, apolipoprotein E, albumin and serotransferrin using peptide standards (GenScript, UK). Peptide sequences and their respective ion transitions monitored in the ChP SILK assay for unlabeled and labeled proteins are provided in Table S3.

Samples were injected (0.5 µL) onto an Acquity Premier peptide BEH C18 column (300 Å, 1.7 μm, 2.1 × 50 mm) held at 50°C and peptides were separated over a 16-minute reverse-phase UPLC gradient. Details of the UPLC gradient and mass spectrometer parameters are further detailed in Leckey et al. (*54*).

Acquired data was imported into Skyline for peak picking and peak integration. Peak areas were exported as a .csv file ready for further statistical analysis.

#### Compartmental modelling of lumbar vs ventricular CSF

To determine whether differences in clearance rates between NPH and SAH-control participants could be attributed to CSF compartmental differences (lumbar vs ventricular sampling respectively), lumbar CSF SILK data previously studied at Washington University in St Louis (Control [L] cohort) was compared to ventricular CSF SILK data (Control [V] cohort) for transthyretin during modelling. The Control (L) cohort included cognitively normal individuals who were amyloid negative on amyloid PET or CSF Ab42/40 ratio. A detailed SILK protocol for these five subjects has been previously described (*14*). Briefly, study participants were administered an initial bolus of ^13^C_6_-leucine for 10 minutes of 3 mg/kg prior to continuous intravenous infusion at a rate of 1.8 - 2.5 mg/kg/hr for 9 hours. CSF and plasma samples were collected at 1- or 2hr intervals over a total period of 36hr and enrichment of ^13^C_6_-leucine was quantified using capillary gas chromatography-mass spectrometry (GC-MS). Stable isotope labeling kinetics data for the validation control subjects was acquired by untargeted high-resolution mass spectrometry at Université de Montpellier using the SILAV approach as previously described (*55*).

### Quantitation and statistical analysis

#### Quantitation of ChP kinetic rates in vitro *and* in vivo

The fractional synthesis rate (FSR) of each protein was calculated from the linear regression of the upslope during the chase divided by the mole fraction labeled leucine (^13^C_6_-leucine) at plateau in plasma during the pulse. Due to NPH study participants awaiting a surgical theatre slot, the timing of which could be unpredictable, pulse duration varied from participant to participant, thus FSRs for the NPH cohort were calculated using the plasma leucine TTR from a subject who reached a steady plateau of the tracer during labeling. Fractional clearance rate (FCR) was derived from the negative slope of the natural logarithm of the clearance portion of the kinetic curve (from peak of labeling onwards). FCRs were captured in 8/10 NPH participants due to difference in chase periods between participants.

Protein half-life was calculated using the exponential decay equation *t* 1/2 = ln(2)/*k*, where *k* is calculated from the non-linear, exponential regression of the clearance portion of the kinetic curve.

To determine if FSR and FCR measures were significantly different between controls and NPH participants, Mann-Whitney U tests were performed in IBM SPSS Statistics (version 27, IBM). Figures displaying the labeled/unlabeled peptide percentages over time were generated using Python.

#### Statistics

All statistical analyses were performed using Excel, Prism v.9.5.1 (GraphPad Software), IBM SPSS Statistics (version 27, IBM), R (4.4.2) or Python software.

To assess data normality, Shapiro-Wilk tests were performed. Spearman’s correlation analyses were used to assess the association between CSF production rate measured in-clinic by LiquoGuard7 and FSR of transthyretin by SILK *in vivo*. Unpaired t-test was performed to assess if there were significant differences in TTHY half-life between NPH patients and controls. p ≤ 0.05 was considered statistically significant.

## Acknowledgements

We are grateful to the participants in this study, and their families, and to the Leonard Wolfson Experimental Neurology Centre for facilitating sample processing.

## Funding

RWP is the recipient of an Alzheimer’s Association Clinician Scientist fellowship AACSF-20-685780, Alzheimer’s disease Part the Cloud award PTC-22-982162 and a Rosetrees Race Against Dementia Team award supported by the Q Charitable Trust. He is supported by the UCLH Queen Square BRC. TAG was supported by research funding from the Alzheimer’s Association (23AARFD-1029918).

## Author Contributions

Conceptualization: RWP

Methodology: RWP/KM

Investigation: RWP/ECM/CAL/TAG/AT/LT/LW

Project administration: SB/ECM/RWP

Supervision: RWP/RJBKM

Funding acquisition: RWP

Data analysis: CAL/TAG/ECM/MSS/MS/WC/DE/SL/JV/SW/RJB/KM/DLE/LP

Data collection: RWP/EM/KT/MS/AT/LW/LT/LP

Ethical approval: RWP/SB

Writing – original draft: RWP/CAL/TAG/ECM/LP

Writing – review & editing: RWP/CAL/TAG/ECM/EM/KT/AA/MSS/MS/WC/SB/DE/AT/LW/LT/SL/JV/SW/RJB/KM/DLE/LP

## Competing Interests

KM is a shareholder of Guilford Street Laboratories Ltd. RJB has received research funding from Avid Radiopharmaceuticals, Janssen, Roche/Genentech, Eli Lilly, Eisai, Biogen, AbbVie, Bristol Myers Squibb, and Novartis. Washington University and RJB have equity ownership interest in C2N Diagnostics and receive income based on technology (stable isotope labeling kinetics, blood plasma assay, methods of diagnosing AD with phosphorylation changes, neurofilament light chain assays and materials) licensed by Washington University to C2N Diagnostics. RJB receives income from C2N Diagnostics for serving on the scientific advisory board. RJB serves as an unpaid member on scientific advisory boards for Roche and Biogen.

## Data and materials availability

Data available from the corresponding author on reasonable request.

## Notes

### Author Declarations

The NHS Health Research Authority, Research Ethics Committee London-Bloomsbury gave ethical approval for the UCL NPH SILK study. All individuals provided informed written consent. Individuals with suspected idiopathic normal pressure hydrocephalus were recruited from the specialist hydrocephalus service at the National Hospital for Neurology and Neurosurgery, Queen Square.

