## Supplementary Information for "Cerebrospinal fluid clearance impairment captured using stable isotope labeling kinetics (SILK) in normal pressure hydrocephalus"

**A**

**B**

**C**

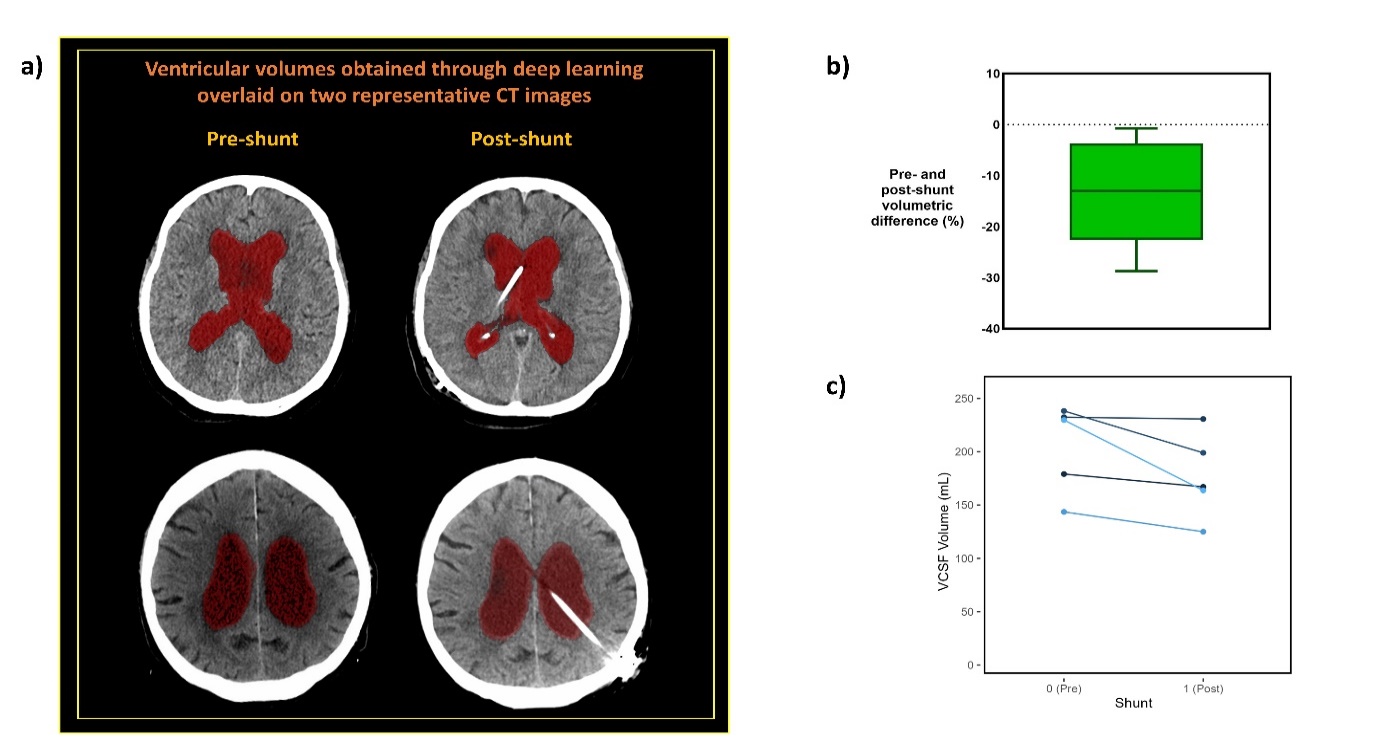

**Figure S1. Summary of NPH imaging:** (**A**) VCSF segmentation outputs obtained using deep-learning-derived CT brain quantification pipeline in two representative pre- and post-shunt paired CT datasets. (**B**) VCSF volume changes (in %) between pre- and post-shunt VCSF images. (**C**) VCSF volumes in millilitres before and after shunt. VCSF, Ventricular cerebrospinal fluid.

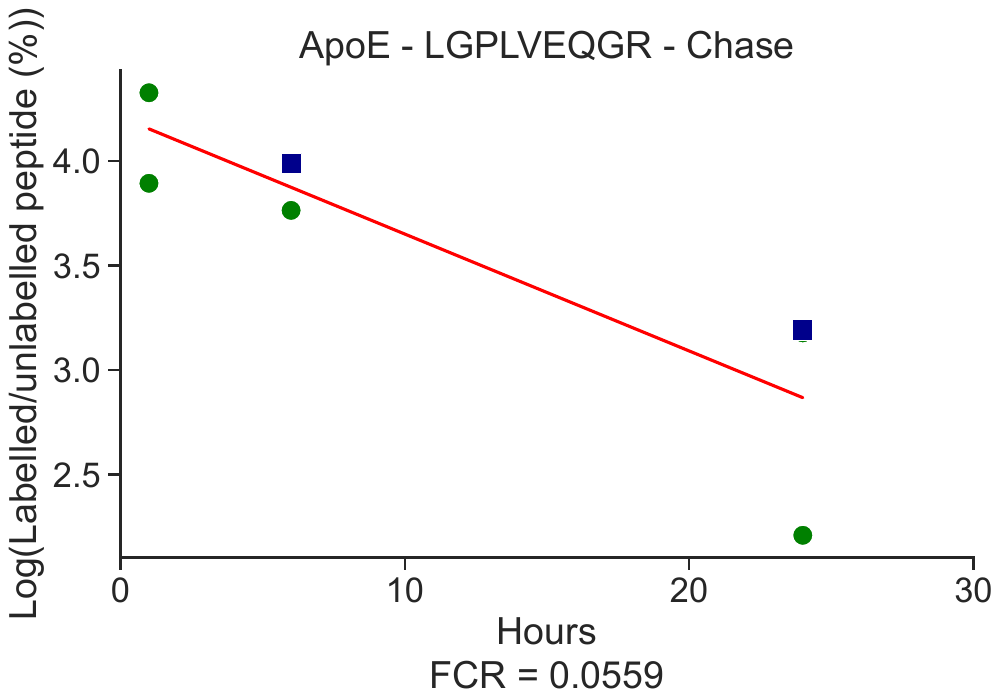

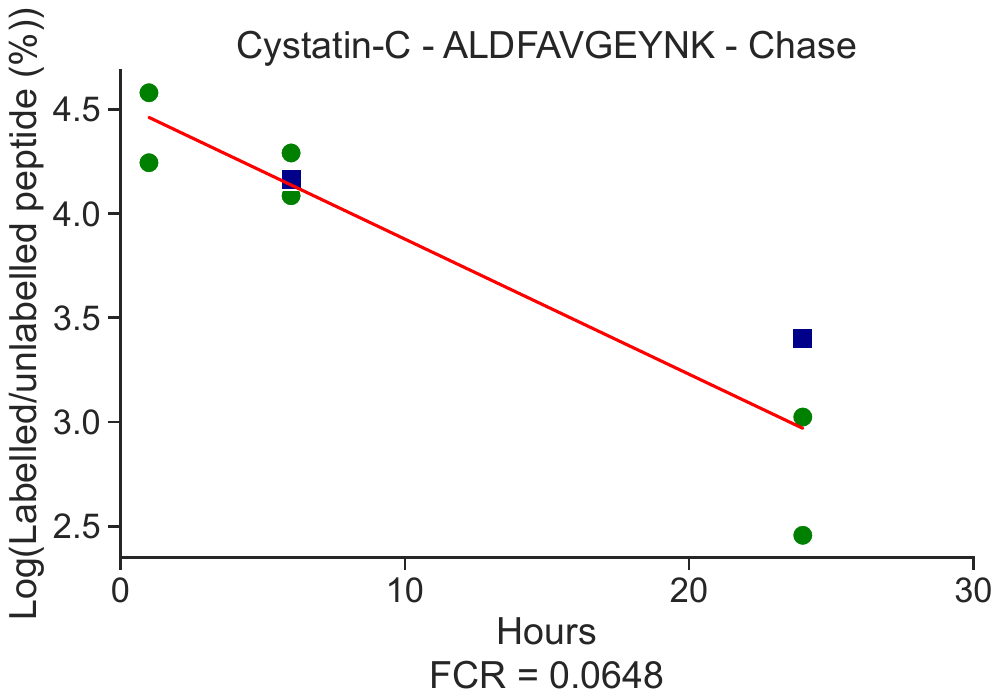

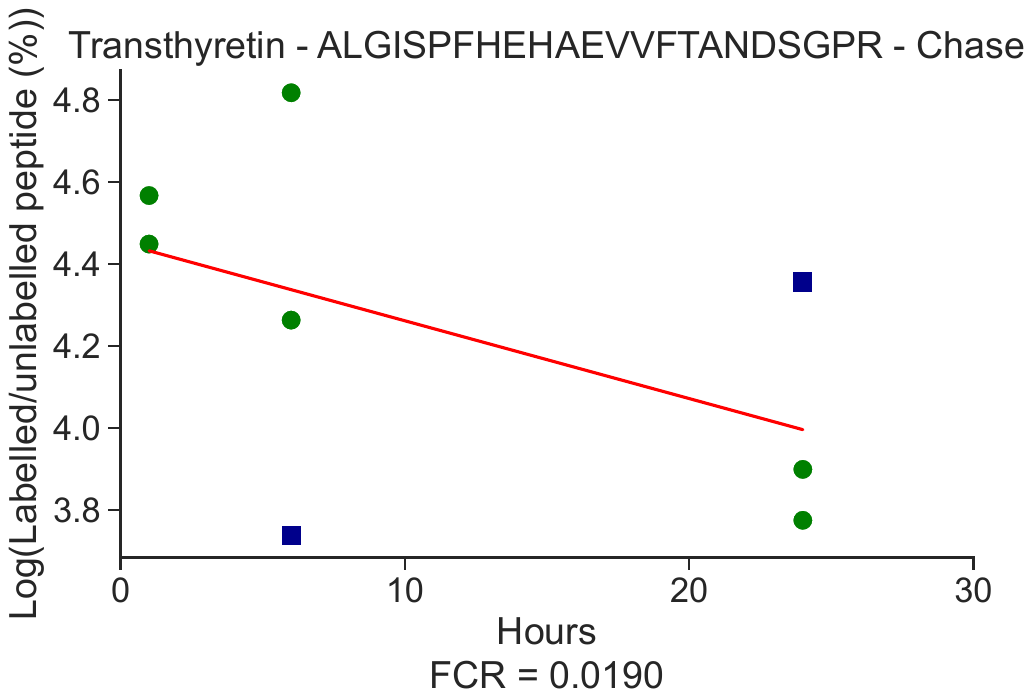

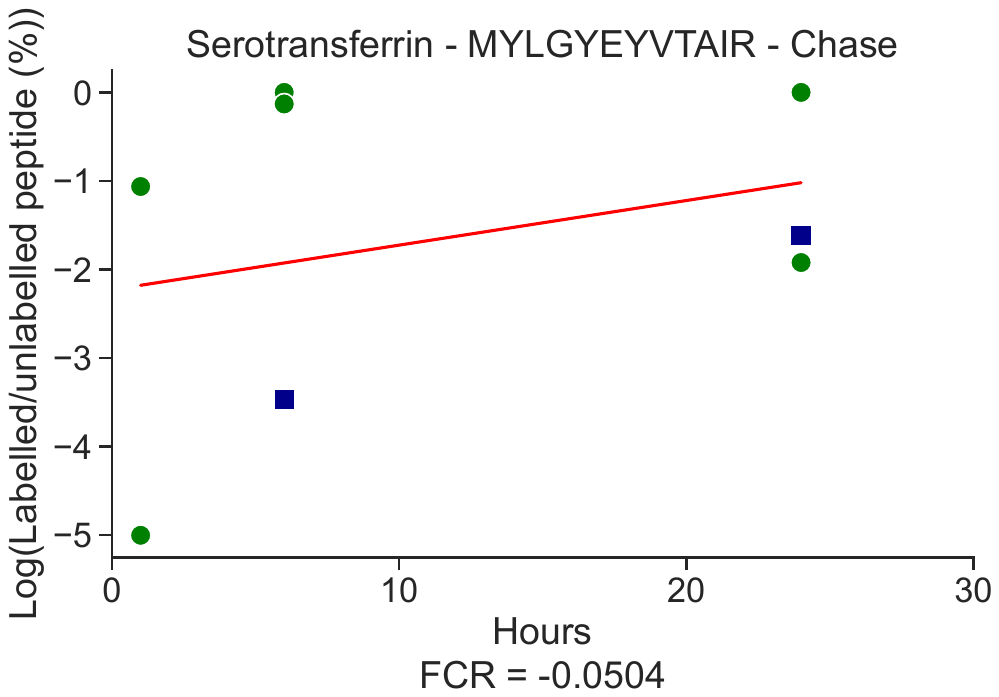

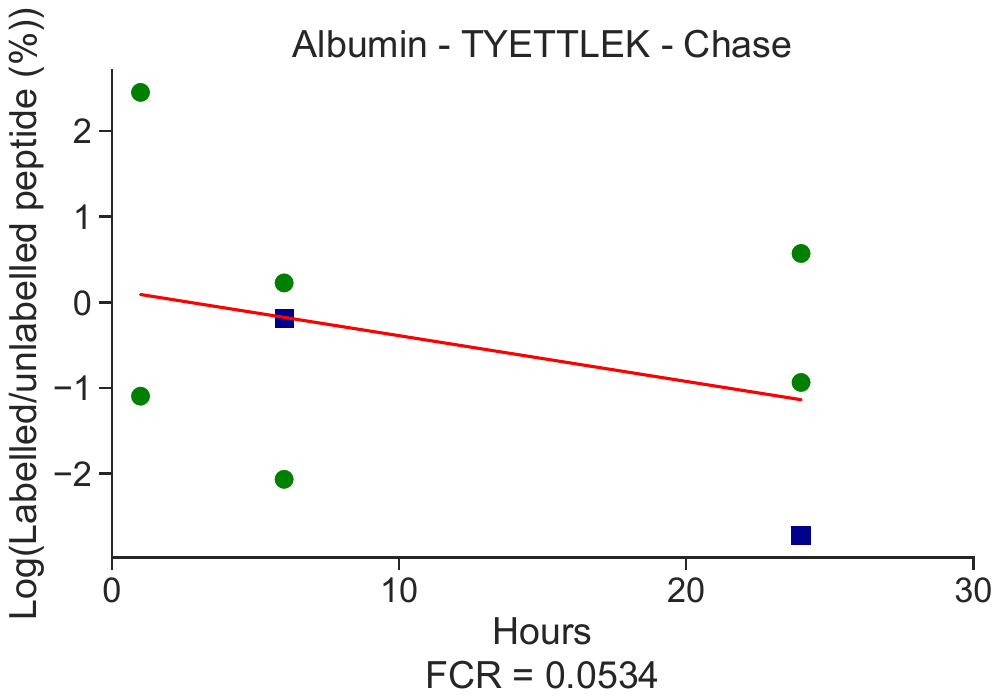

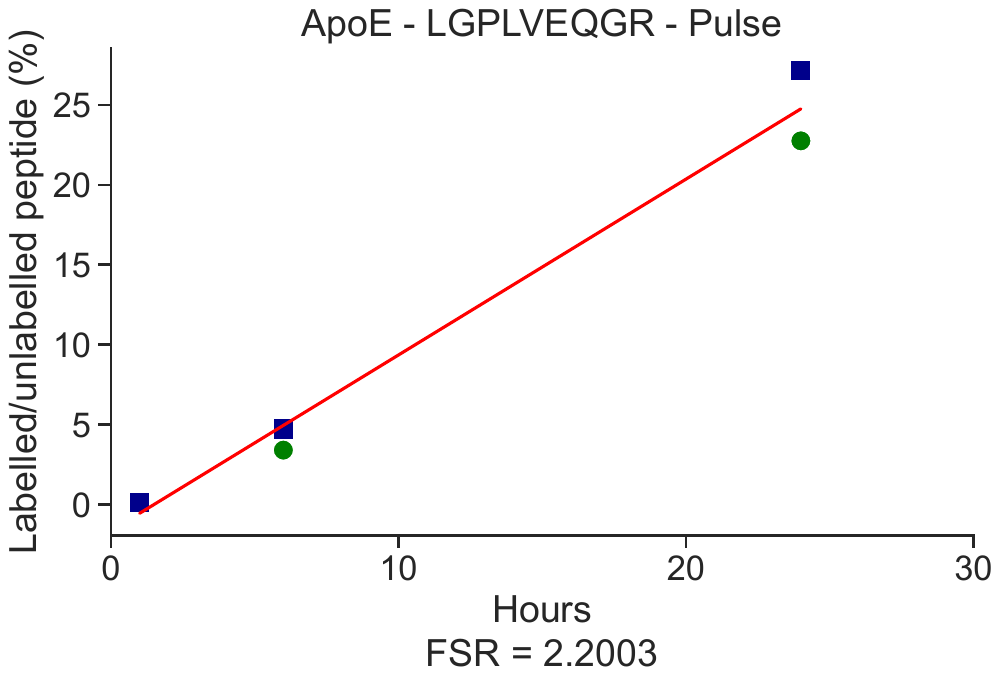

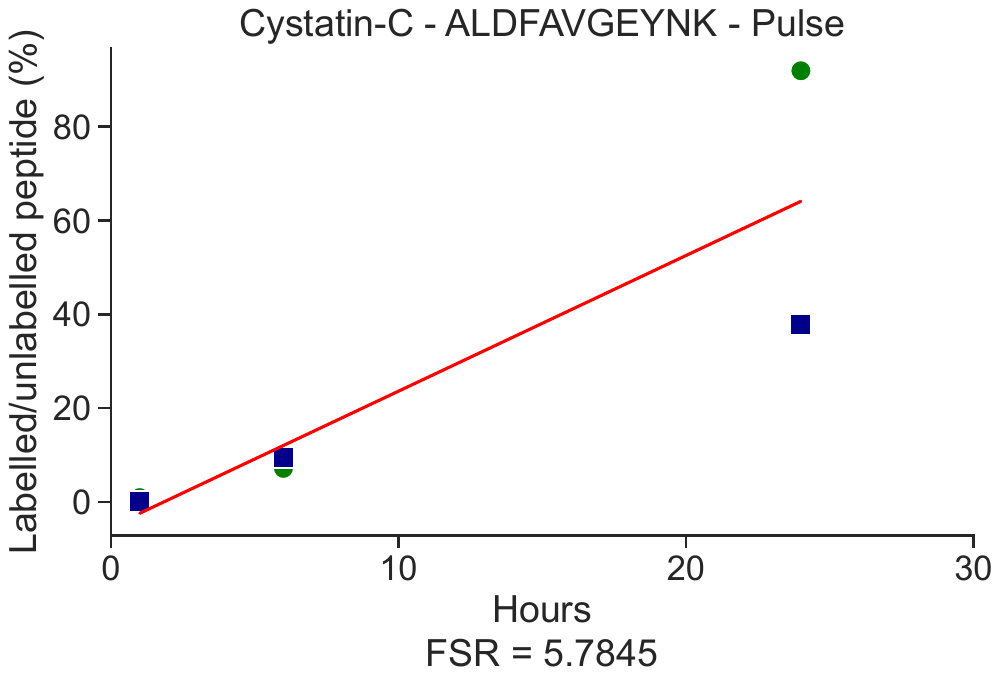

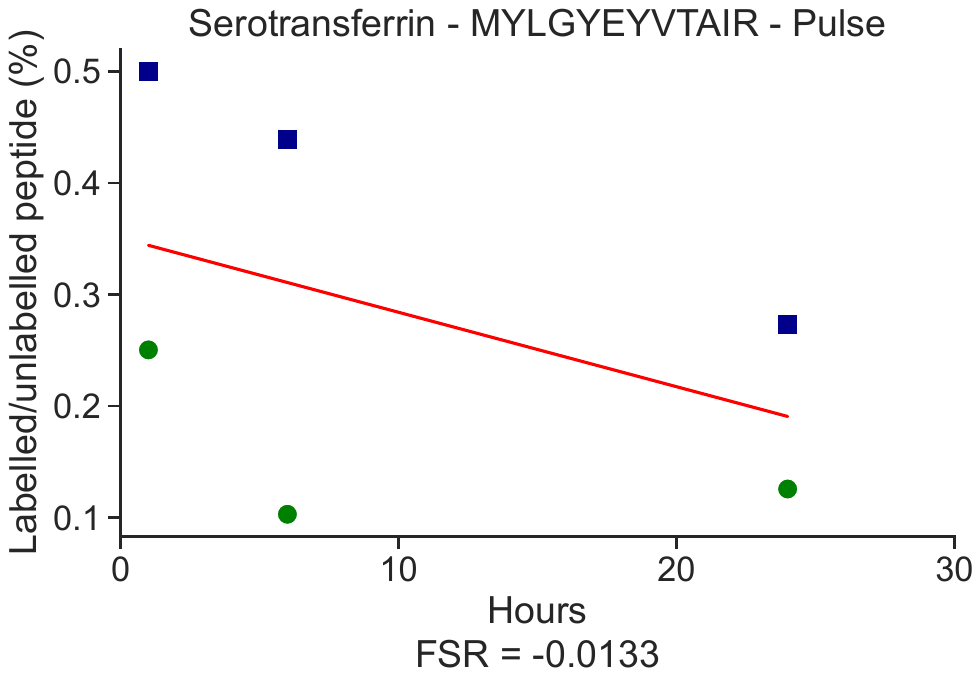

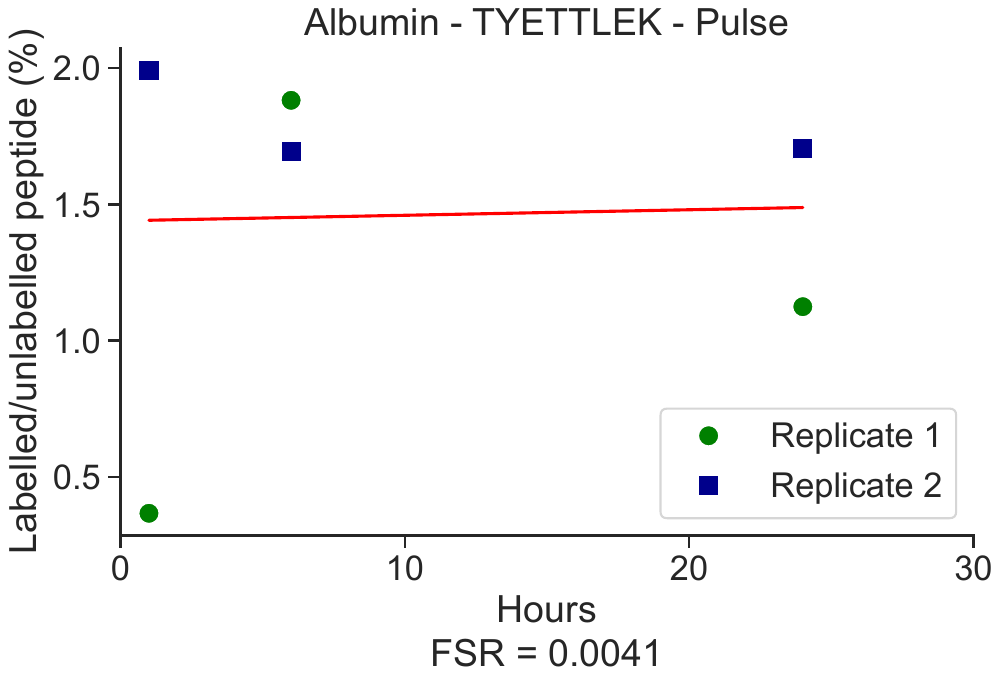

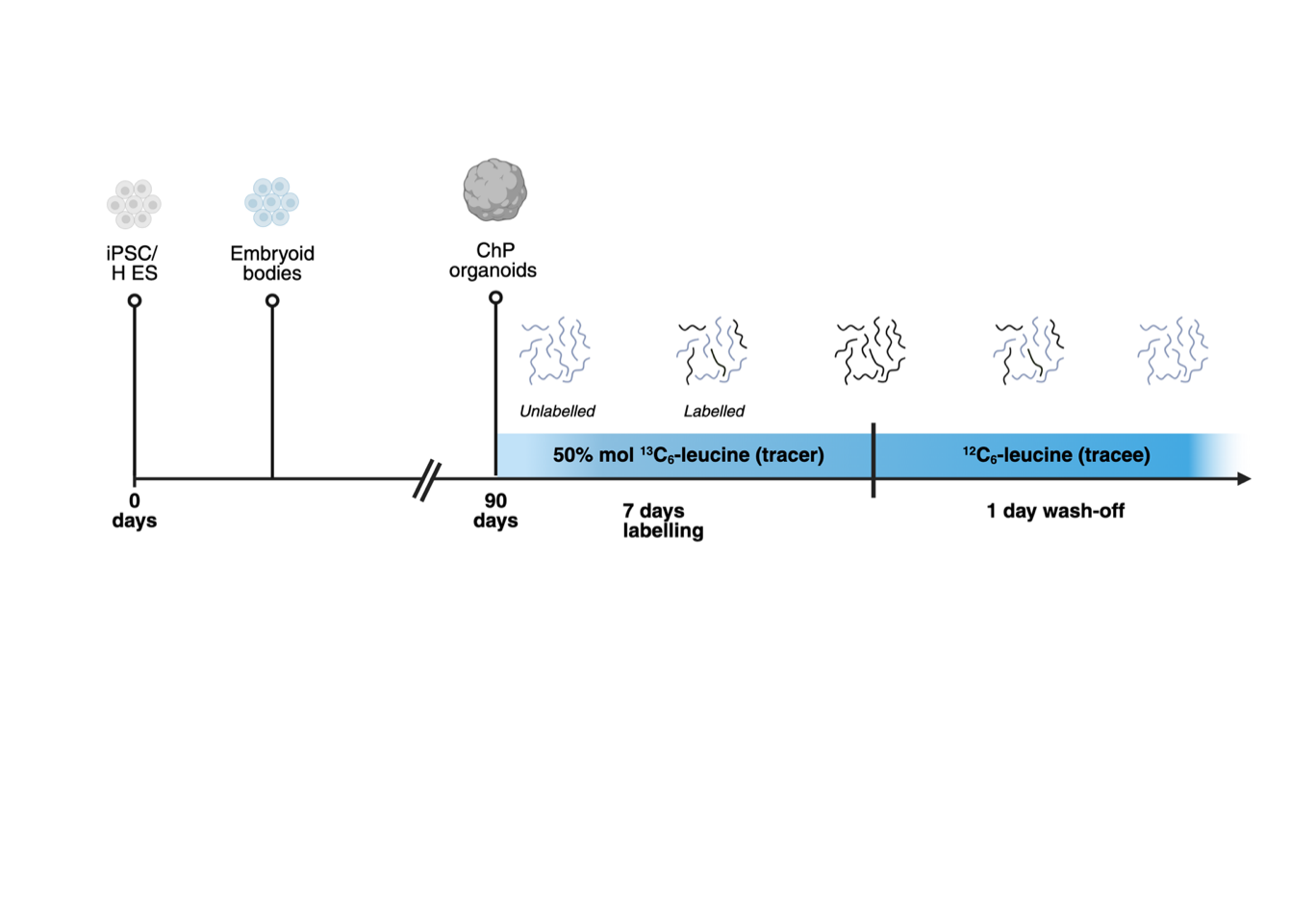

**K**

**J**

**I**

**H**

**G**

**F**

**E**

**D**

**C**

**B**

**A**

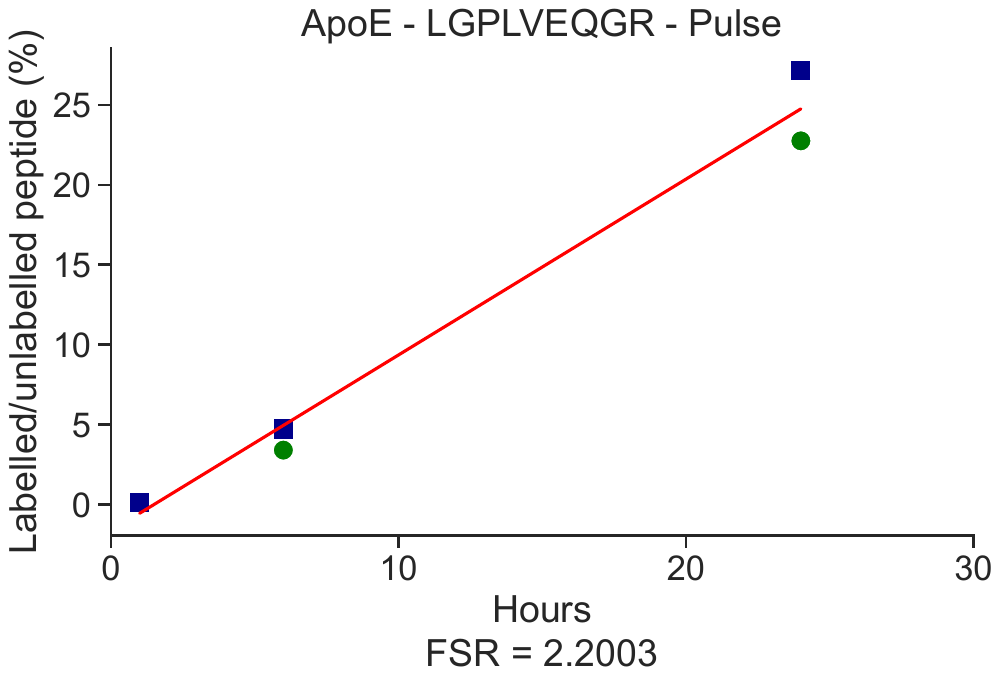

f

g

h

i

j

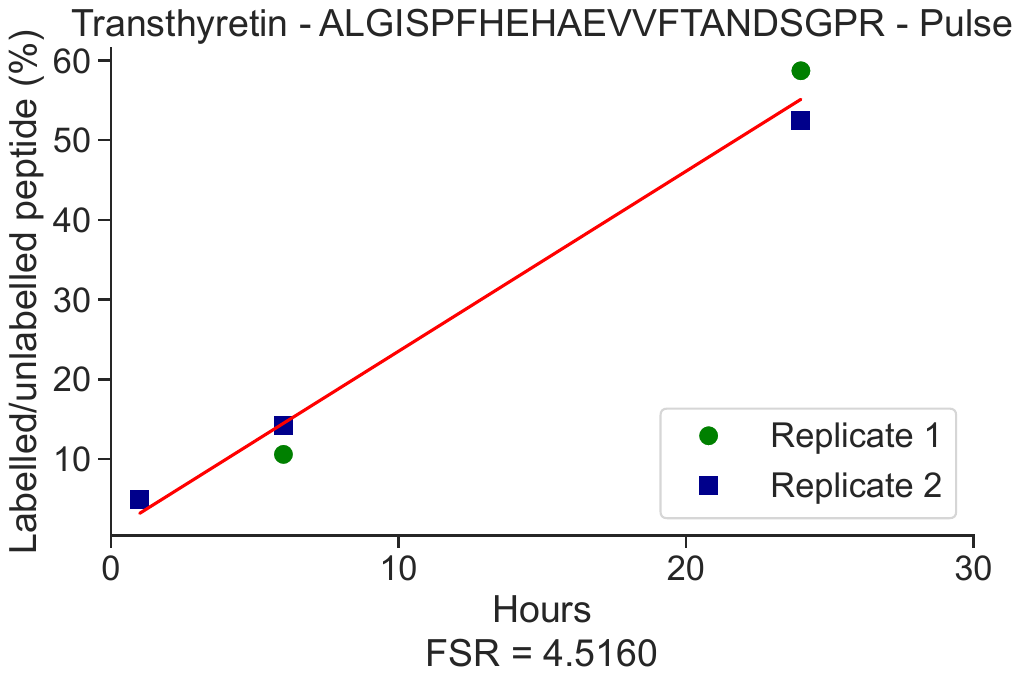

**Figure S2. Choroid plexus protein kinetics in ChP organoid CSF (iCSF). Fractional synthesis rates of plasma and choroid plexus derived proteins in choroid plexus organoid CSF (iCSF):** (**A**) Schematic showing development and ^13^C_6_-leucine labeling of ChP organoids. (**B**) Albumin. (**C**) Serotransferrin. (**D**) TTHY. (**E**) Cys-C. (**F**) ApoE. Fractional clearance rates of plasma and ChP derived proteins in ChP organoid lysate: (**G**) Albumin. (**H**) Serotransferrin. (**I**) TTHY. (**J**) Cys-C. (**K**) ApoE. ApoE, apolipoprotein E; ChP, choroid plexus; Cys-C, cystatin C; TTHY, transthyretin.

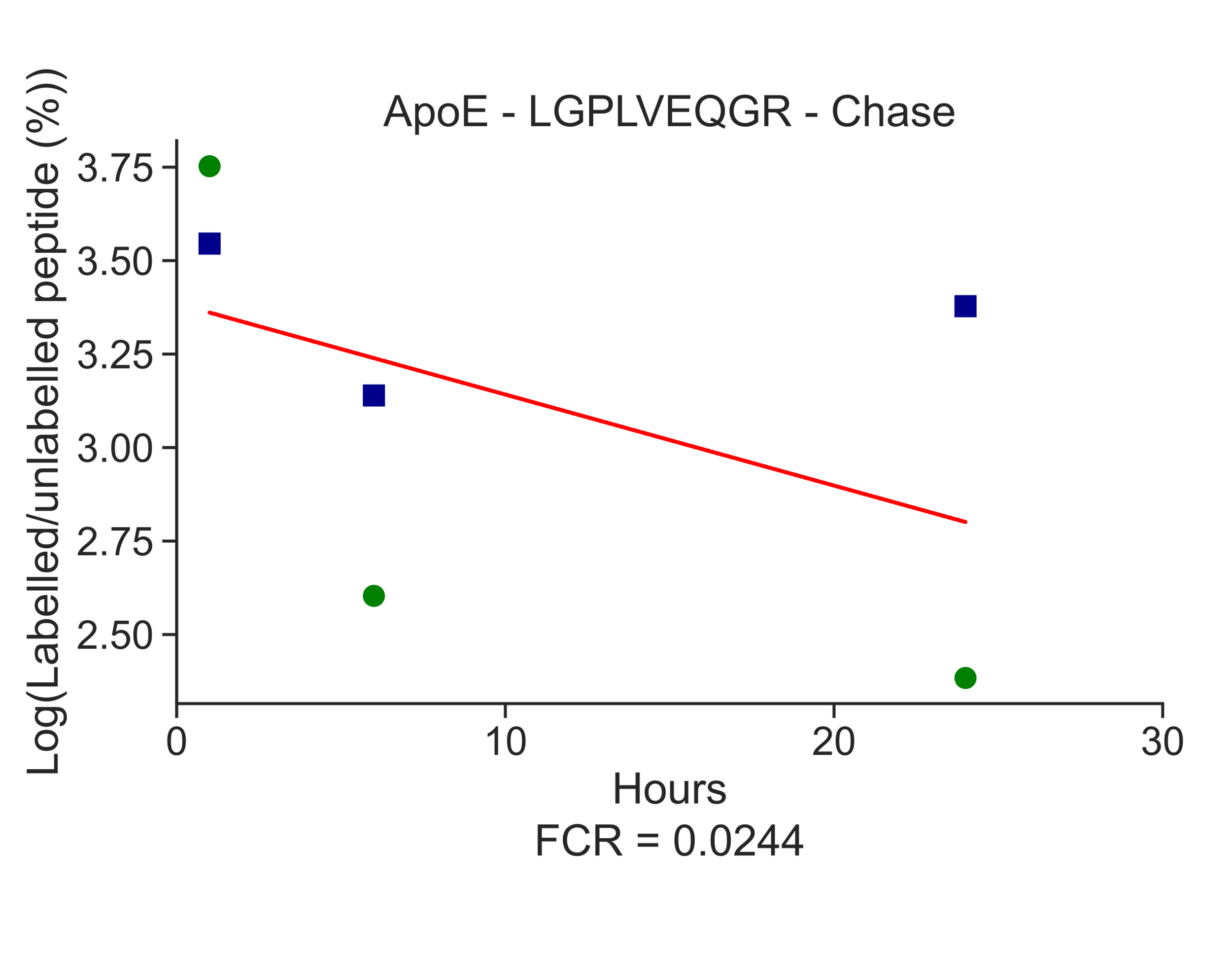

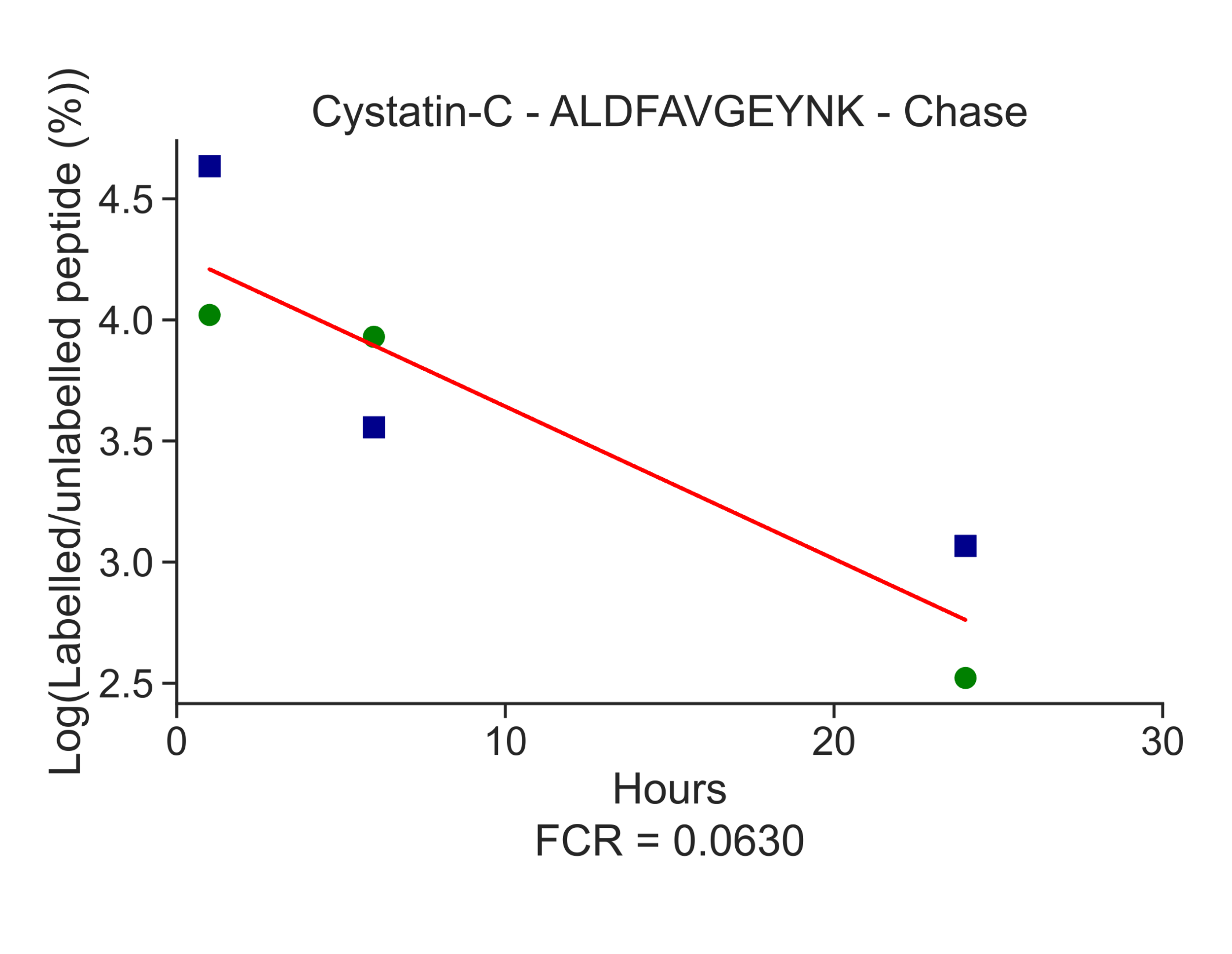

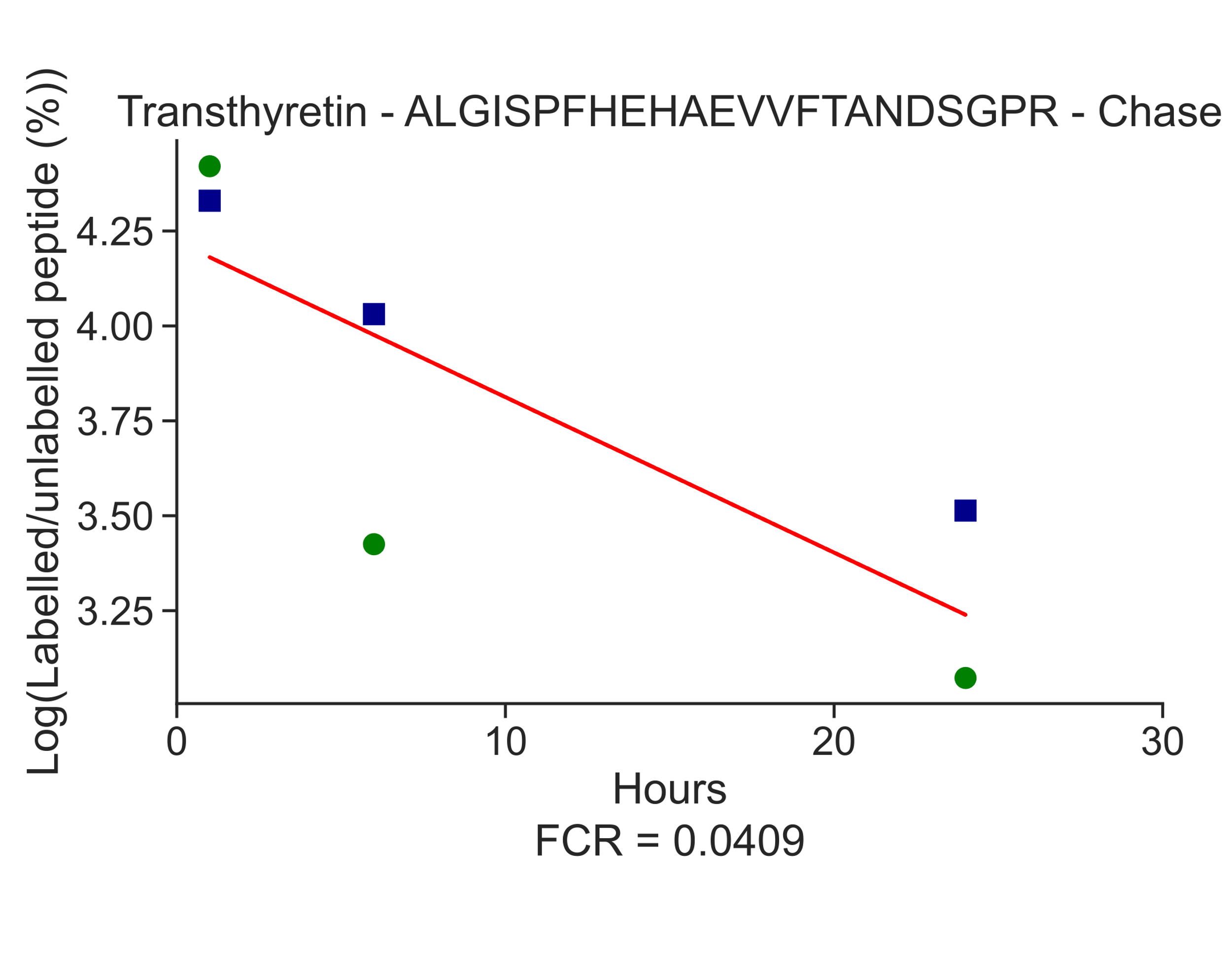

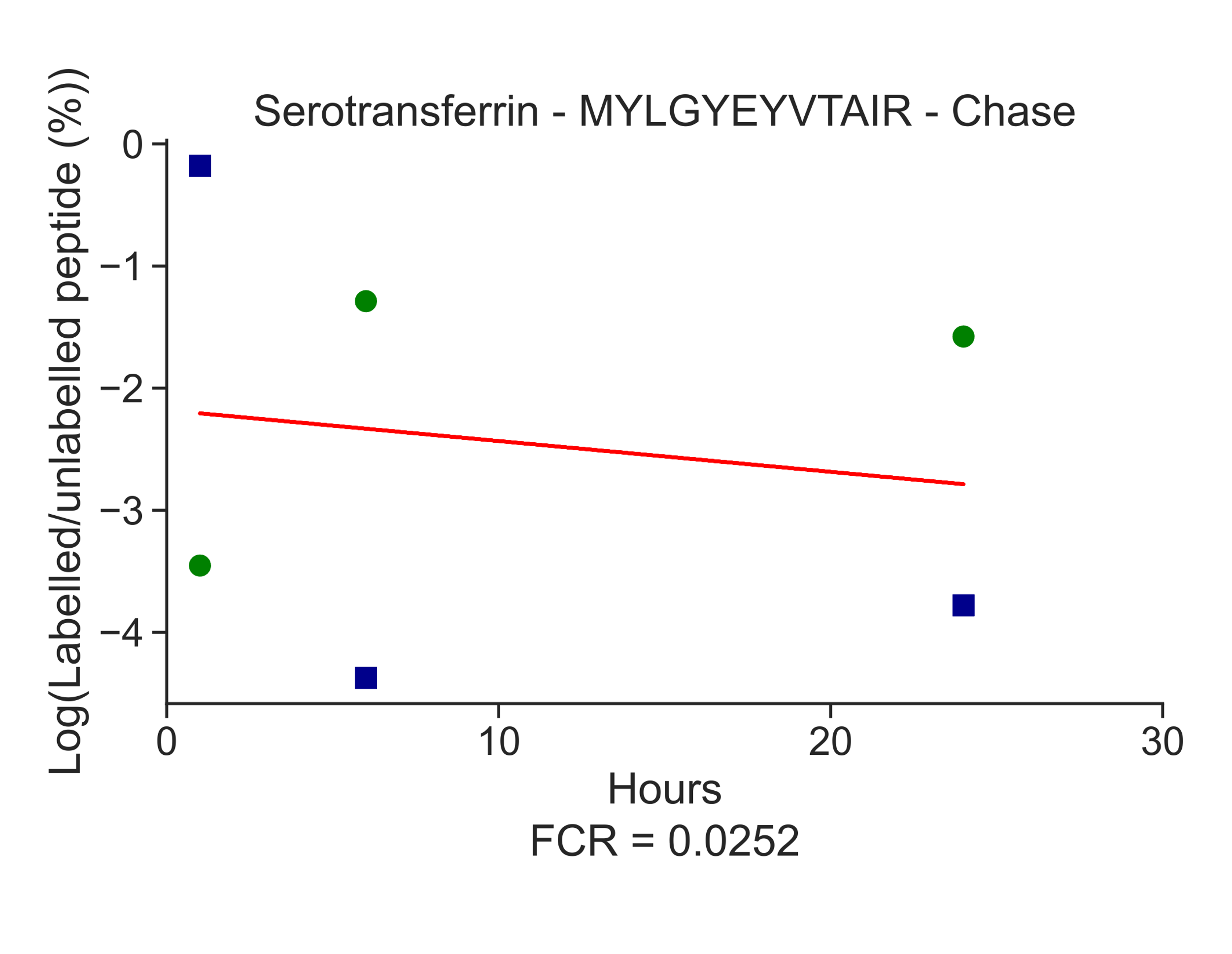

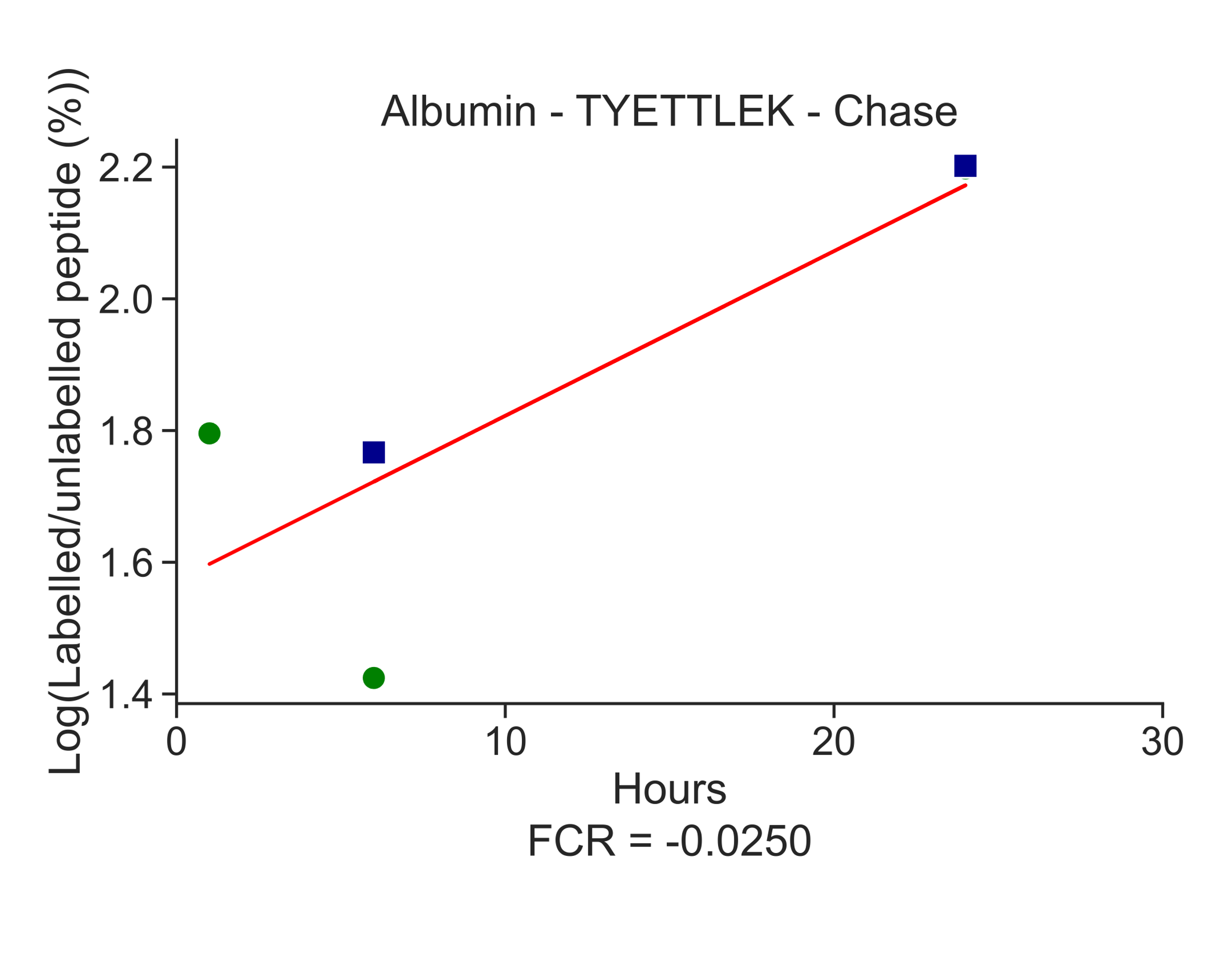

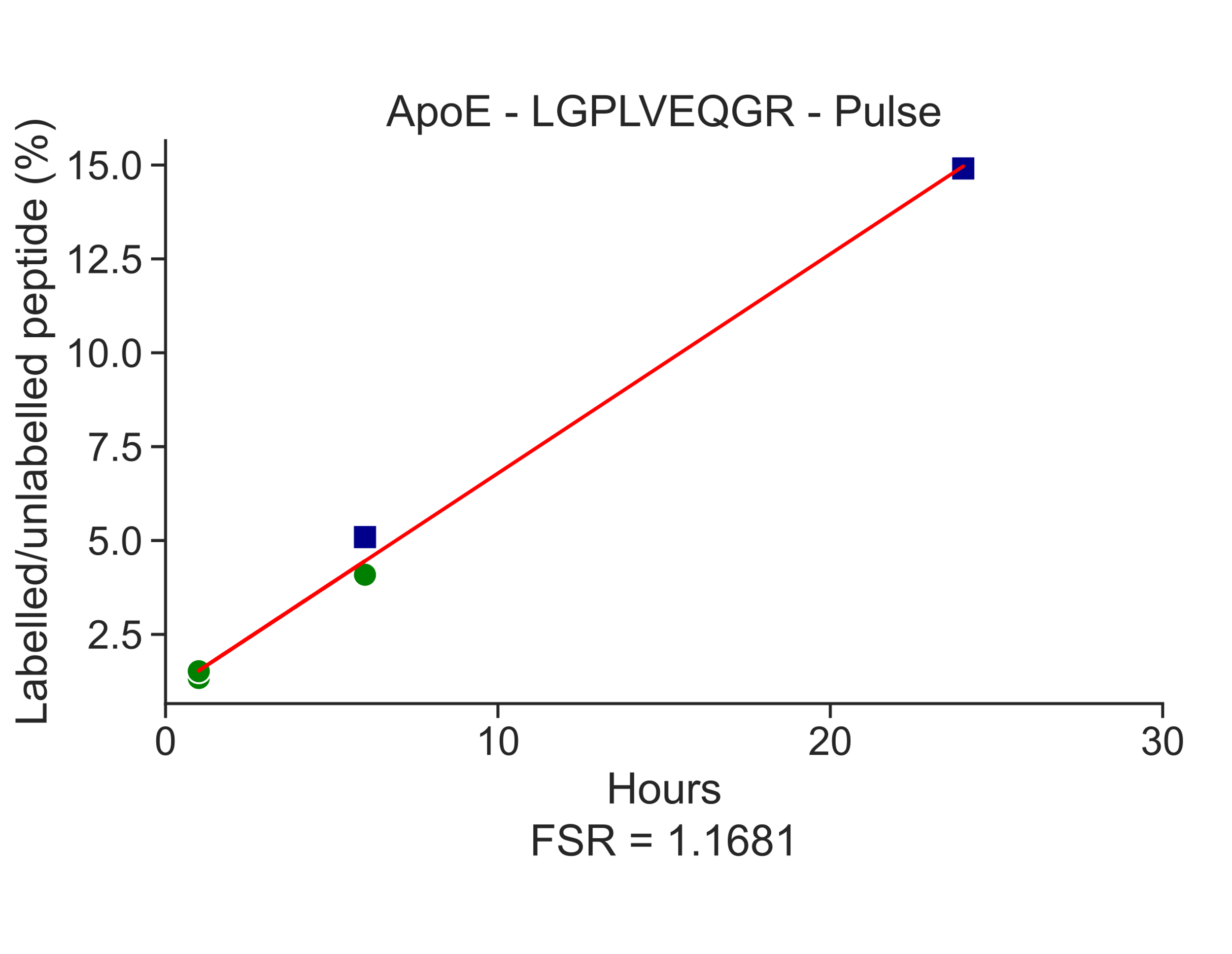

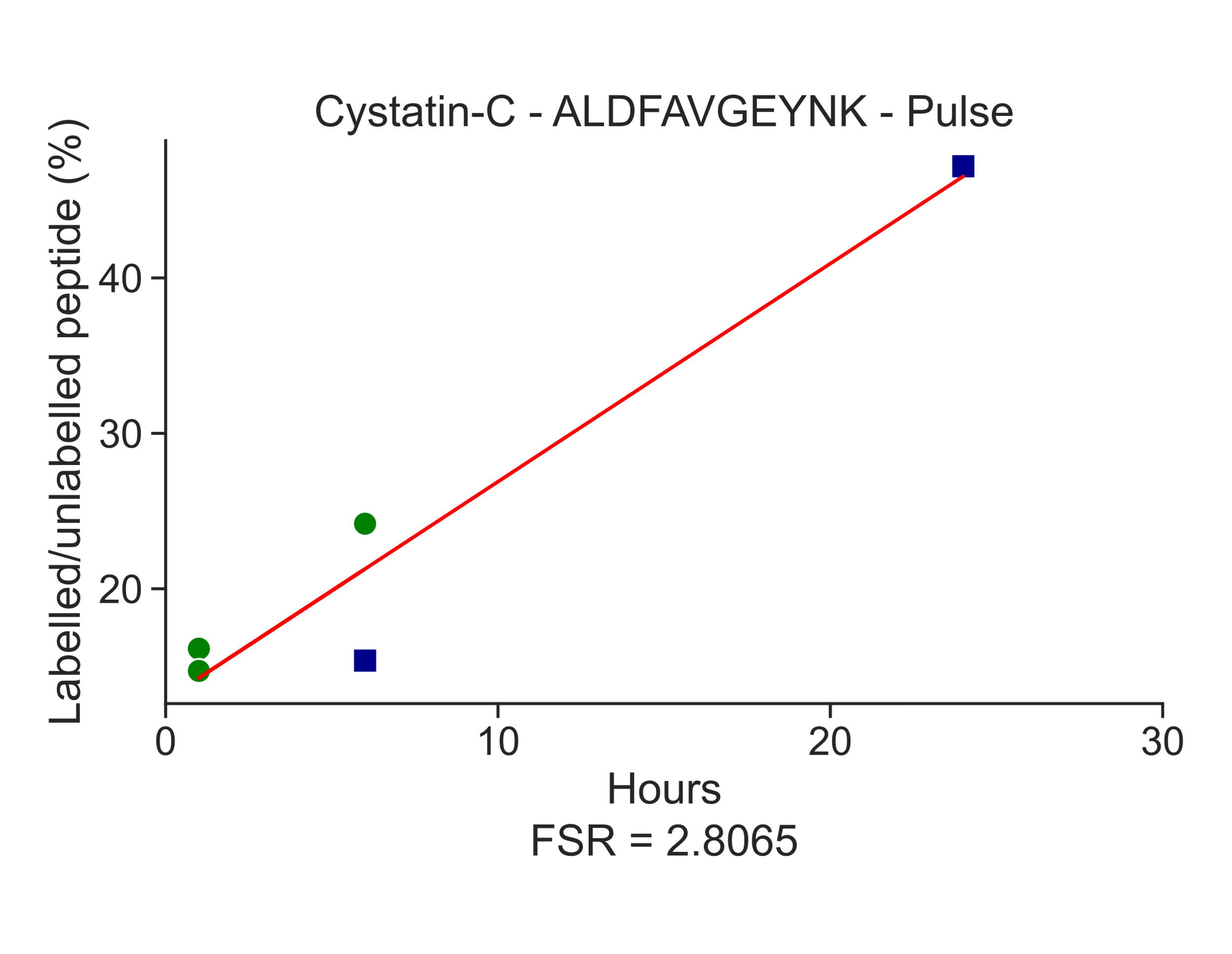

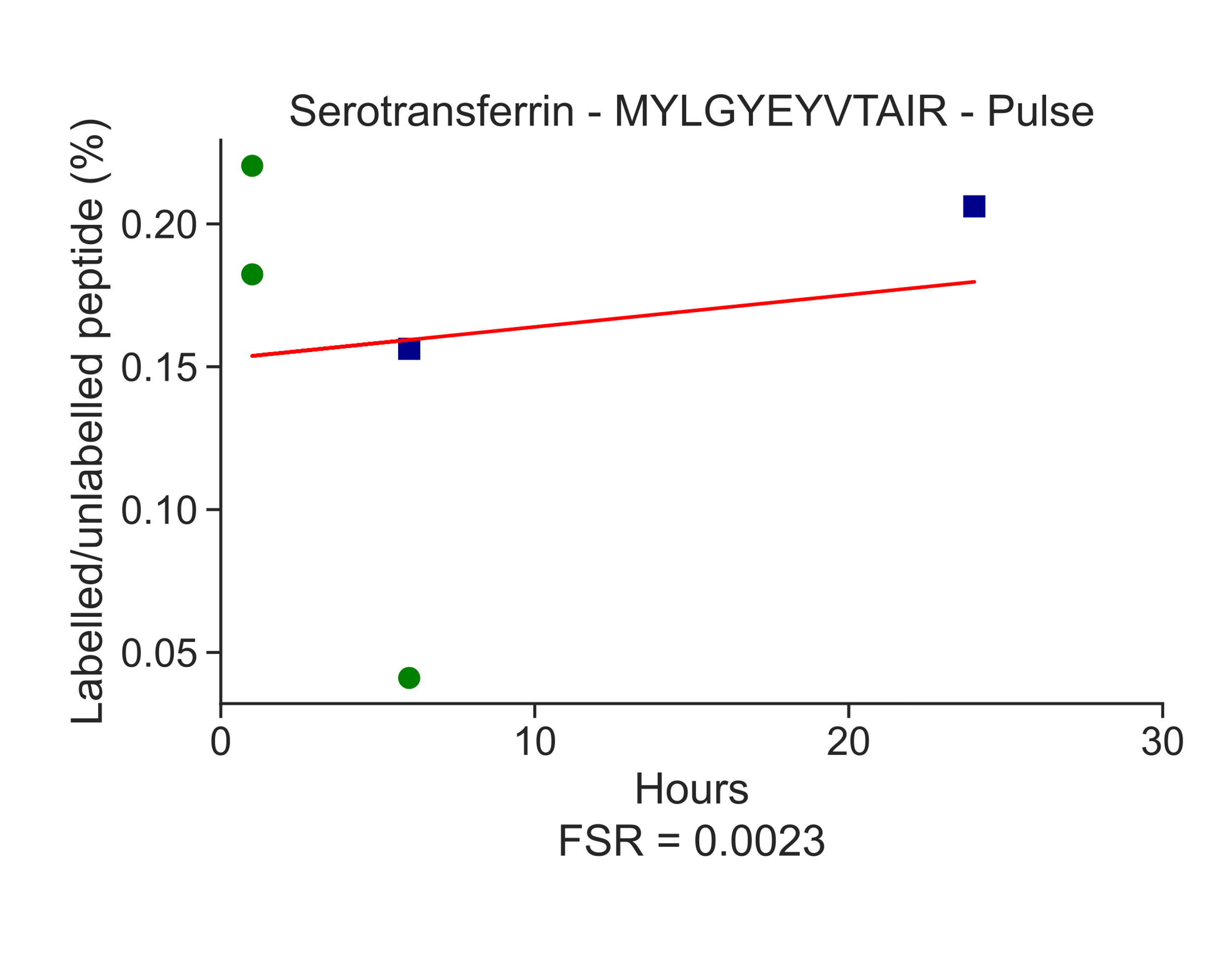

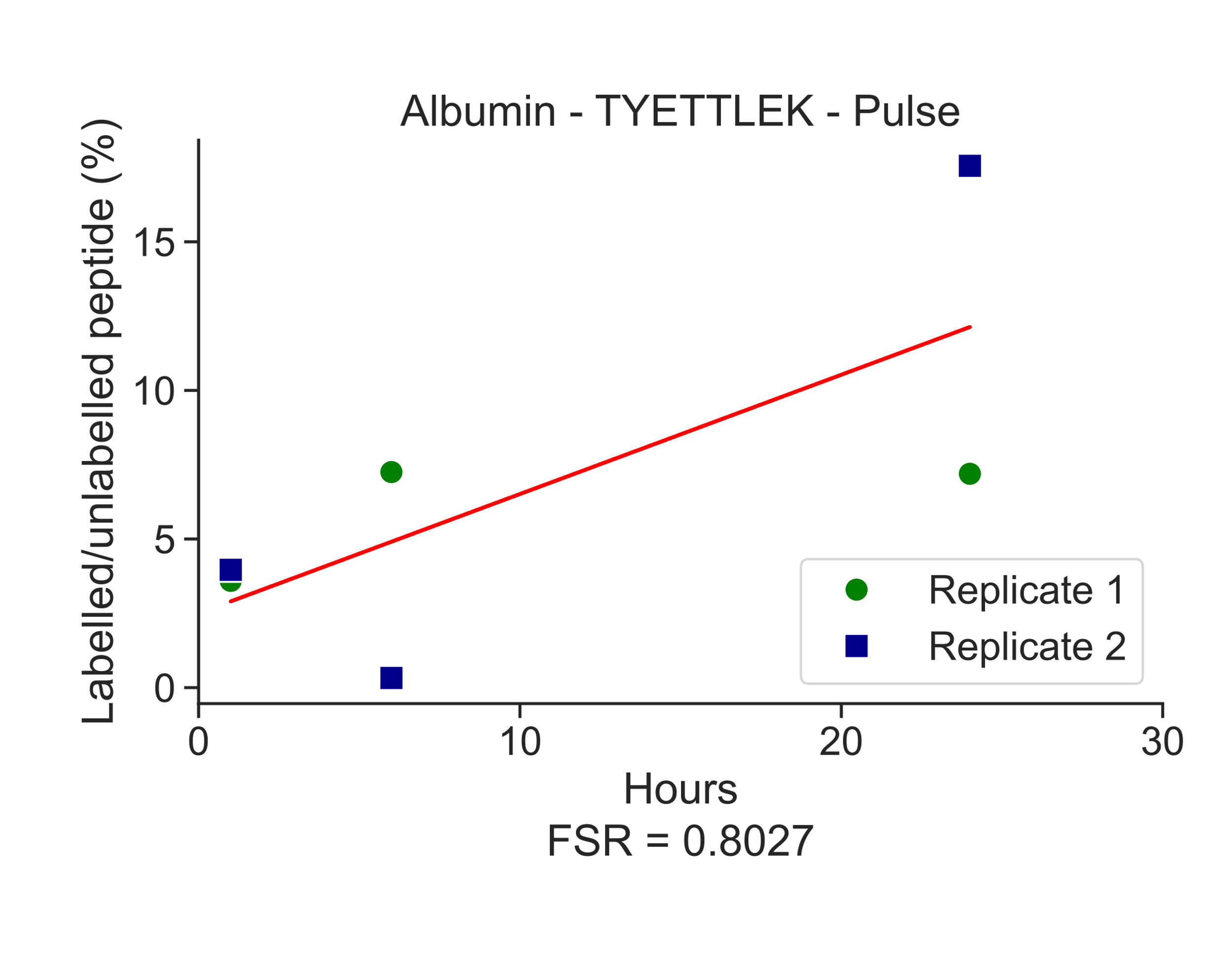

**J**

**I**

**H**

**G**

**F**

**E**

**D**

**C**

**B**

**A**

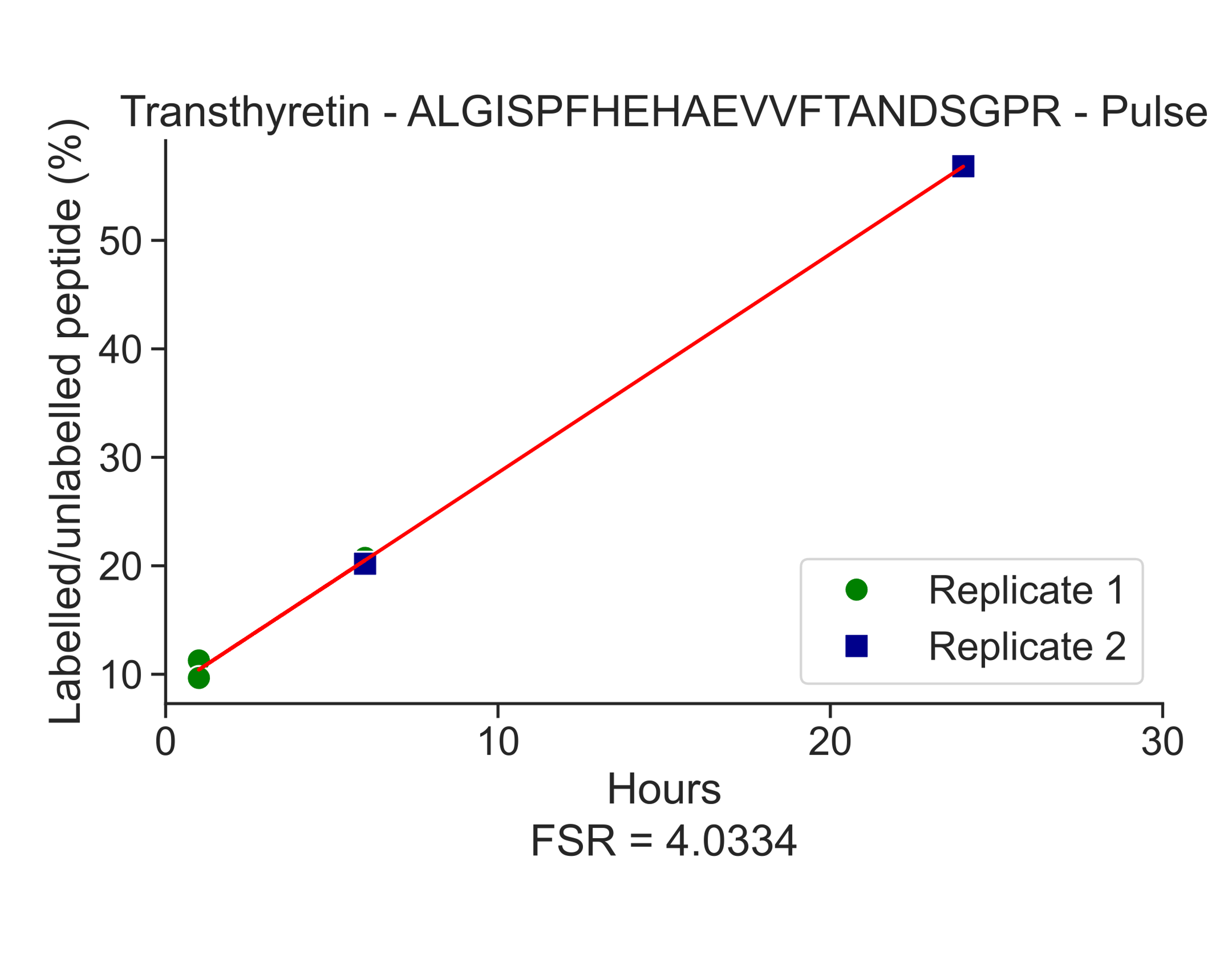

**Figure S3. Choroid plexus protein kinetics in ChP organoid lysates. Fractional synthesis rates of plasma and choroid plexus derived proteins in choroid plexus organoid lysate:** (**A**) Albumin. (**B**) Serotransferrin. (**C**) TTHY. (**D**) Cys-C. (**E**) ApoE. Fractional clearance rates of plasma and ChP derived proteins in ChP organoid lysate: (**F**) Albumin. (**G**) Serotransferrin. (**H**) TTHY. (**I**) Cys-C. (**J**) ApoE. ApoE, apolipoprotein E; ChP, choroid plexus; Cys-C, cystatin C; TTHY, transthyretin.

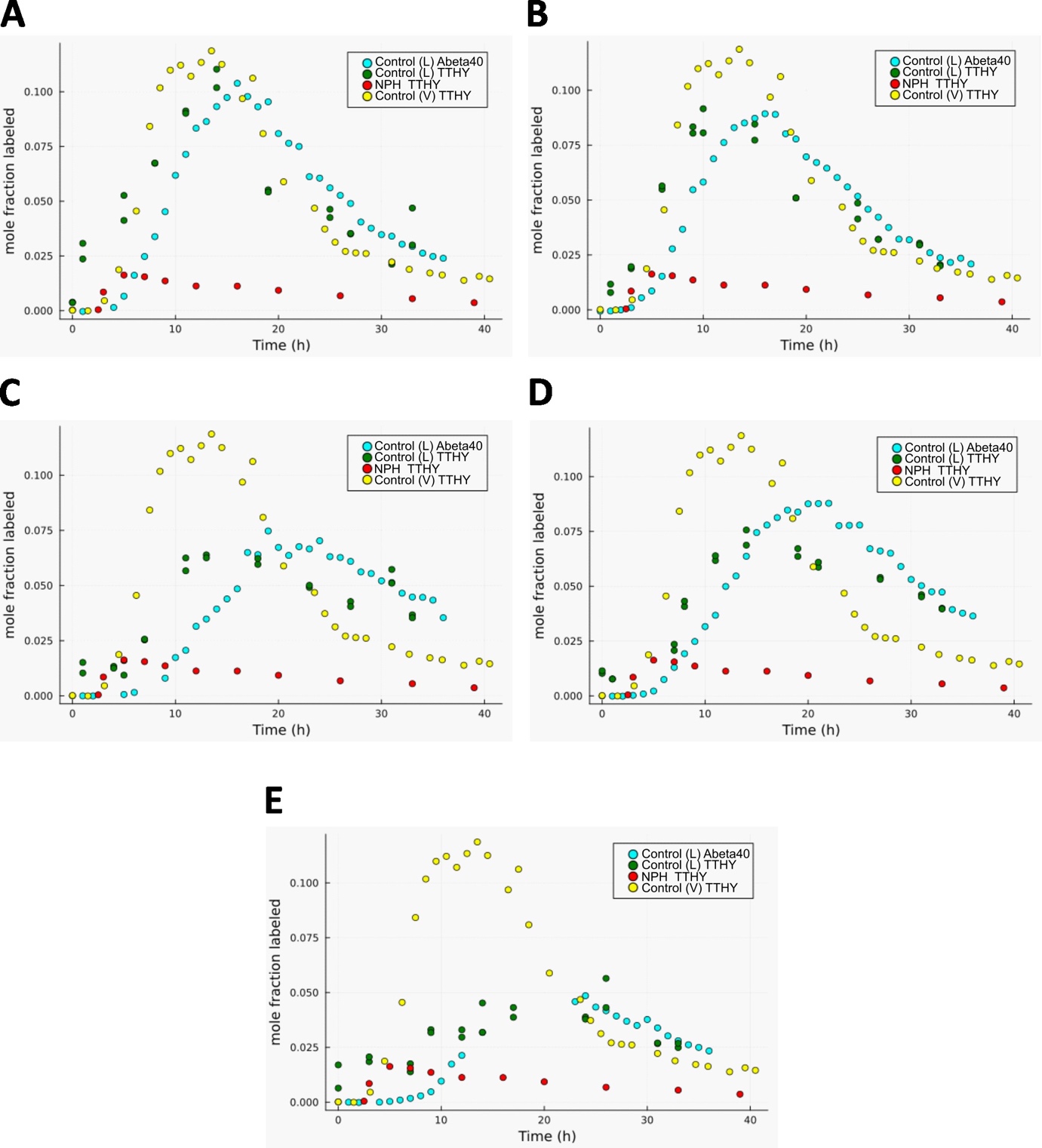

**Figure S4. Comparison of protein kinetics between five lumbar CSF control subjects.** (**A-E**) Kinetics of amyloid beta 40 peptide (Abeta40) and transthyretin (TTHY) of each subject. For aiding comparisons, the average turnover of TTHY in the NPH- and pSAH (ventricular CSF) cohorts are shown. Abeta, amyloid beta 40 peptide; L, lumbar; NPH, normal pressure hydrocephalus; TTHY, transthyretin; V, ventricular.

**Supplementary Tables**

| **Table S1 \| Demographic and biomarker data for NPH and control cohorts.** | | | |
| --- | --- | --- | --- |
|  | NPH  (*n* = 10) | Controls (post SAH)  (*n* = 4) | Controls (validation)  (n=5) |
| Age at LP (years) | 75 (71-78) | 57 (46-65) | 70 (63-84) |
| % Male | 80 | 0 | 40 |
| % Caucasian | 90 | NA | 80 |
| MMSE | 28 (26-28)  (*n* = 9) | NA | NA |
| Baseline 10m walk time (seconds) | 11 (10-23)  (*n* = 9) | NA | NA |
| Number responding to shunting | 7 | NA | NA |
| % Executive Dysfunction | 50 | NA | NA |
| % Episodic memory problems | 70 | NA | NA |
| % Language Impairment | 20 | NA | NA |
| % Gait disturbance | 100 | NA | NA |
| % Parkinsonism | 20 | NA | NA |
| % Positive for Falls | 44  (*n* = 9) | NA | NA |
| % Urinary Incontinence | 80 | NA | NA |
| % Cerebellar signs | 11  (*n* = 9) | NA | NA |
| % Eye movement abnormalities | 33  (*n* = 9) | NA | NA |
| % Supranuclear gaze palsy | 13  (*n* = 8) | NA | NA |
| % Pyramidal signs  CSF production rate (ml/hr)  Clinical CSF Aβ42/40 | 11  (*n* = 9)  84 (62-89)  0.118 (0.082-0.129)  (*n* = 9) | NA  NA  NA | NA |

Median and interquartile ranges are shown

(Interquartile ranges are reported to the nearest whole number)

Where data was missing, the number of subjects for which the data were available is indicated within parentheses

*LP:* lumbar puncture; *MMSE:* mini-mental state examination; *NPH*: Normal pressure hydrocephalus; NA: not available

| **Table S2 \| Summary of ChP peptides, FSR and FCR in NPH and control cohorts.** | | | | | | | | | |
| --- | --- | --- | --- | --- | --- | --- | --- | --- | --- |
| **Protein** | **Peptide Sequence** | **Domain** | **Amino Acids** | **FSR (%/hr)** | | **Sig.** | **FCR (%/hr)** | | **Sig.** |
|  |  |  |  | *NPH* | *Control (V)* |  | *NPH* | *Control (V)* |  |
| Transthyretin | ALGISPFHEHAEVVFTANDSGPR | Transthyretin/ hydroxyisourate hydrolase | 101- 123 | 2.29  (1.27-3.78)  n=8 | 10.79  (9.81-34.10)  n=3 | p < 0.05 (0.012) | 0.74  (0.02-1.16)  n=8 | 7.25  (2.28-11.19)  n=4 | p < 0.005 (0.004) |
|  | TSESGELHGLTTEEEFVEGIYK |  | 69-90 | 1.64  (0.94-2.64)  n=9 | 8.64  (8.36-26.35)  n=3 | p < 0.01 (0.009) | 1.57  (1.05-2.08)  n=8 | 9.45  (5.76-13.79)  n=4 | p < 0.005 (0.004) |
| Cystatin-C | ALDFAVGEYNK | N/A | 52-62 | 0.55  (0.17-1.24)  n=9 | 4.16  (2.23-4.53)  n=3 | p < 0.01 (0.009 | 2.00  (0.74-2.60)  n=6 | 3.45  (2.54-4.36)  n=2 | NS (0.143) |
| Serotransferrin | MYLGYEYVTAIR | Transferrin-like 1 | 332-343 | 0.05  (0.03-0.09)  n=9 | 0.47  (0.27-0.48)  n=3 | p < 0.01 (0.009) | *Not captured* | *Not captured* |  |
| Albumin | TYETTLEK | Albumin 2 | 376-383 | 0.03  (0.01-0.03)  n=8 | 0.10  (0.06-0.12)  n=3 | p < 0.05 (0.012) | *Not captured* | *Not captured* |  |
| Apolipoprotein E | LQAEAFQAR | Lipid-binding and lipoprotein association region | 270-278 | 0.35  (0.08-0.84)  n=8 | 1.77  (1.11-2.00)  n=3 | p < 0.05 (0.012) | 1.40  (0.39-7.58)  n=7 | 1.14  (n=1) | NS (0.827) |
| ***Exploratory analysis between NPH and the Control (L) validation cohort*** | | | | | | | | | |
|  |  |  |  | *NPH* | *Control (L)* |  | *NPH* | *Control (L)* |  |
| Transthyretin | ALGISPFHEHAEVVFTANDSGPR | Transthyretin/ hydroxyisourate hydrolase | 101-123 | 2.29  (1.27-3.78)  n=8 | 4.35  (2.00-7.00)  n=5 | NS (0.093) | 0.74  (0.02-1.16)  n=8 | 4.51  (3.39-6.14) n=5 | p < 0.005  (0.002) |
|  | TSESGELHGLTTEEEFVEGIYK |  | 69-90 | 1.64  (0.94-2.64)  n=9 | 3.95  (0.94-5.03)  n=5 | NS (0.060) | 1.57  (1.05-2.08)  n=8 | 6.22  (2.97-8.50)  n=5 | p < 0.005 (0.004) |

FSR (%/hr) and FCR (%/hr) data represented as group median (min-max values)

NS; Not Significant

Not captured: not long enough chase period to capture clearance curve (only production curve observed)

Sig. (p-values; exact significance) calculated by Mann Whitney U test

Control (V), SAH-Control Cohort (Montpellier); Control (L), Validation lumbar CSF Control Cohort (Washington University in St Louis/Montpellier).

| **Table S3 \| ChP SILK peptides and ion transitions in the multiplexed ChP assay.** | | | | | | |
| --- | --- | --- | --- | --- | --- | --- |
| Protein | Peptide Sequence | Amino Acids | Precursor ion  (m/z) | Precursor charge (z) | Product ion (type) | Product ion  (m/z) |
| Albumin | TYETTLEK | 376-383 | 492.7478 | 2 | y6+ | 720.3774 |
|  |  |  |  |  | y5+ | 591.3348 |
|  | TYETT**L**[**^13^C_6_**]EK |  | 495.7579 | 2 | y6+ | 726.3975 |
|  |  |  |  |  | y5+ | 597.3549 |
| Apolipoprotein E | LQAEAFQAR | 270-278 | 517.2749 | 2 | b3+ | 313.1870 |
|  |  |  |  |  | y3+ | 374.2146 |
|  | **L**[**^13^C_6_**]QAEAFQAR |  | 520.2850 | 2 | b3+ | 313.1870 |
|  |  |  |  |  | y3+ | 374.2146 |
| Cystatin-C | ALDFAVGEYNK | 52-62 | 613.8062 | 2 | y5+ | 610.2831 |
|  |  |  |  |  | b6+ | 617.3293 |
|  | A**L**[**^13^C_6_**] DFAGEYNK |  | 616.8163 | 2 | y5+ | 610.2831 |
|  |  |  |  |  | b6+ | 623.3495 |
| Serotransferrin | MYLGYEYVTAIR | 332-343 | 739.8710 | 2 | y9+ | 1071.5469 |
|  |  |  |  |  | y7+ | 851.4621 |
|  | MY**L**[**^13^C_6_**] GYEYVTAIR |  | 742.8811 | 2 | y9+ | 1071.5469 |
|  |  |  |  |  | y7+ | 851.4621 |
| Transthyretin | ALGISPFHEHAEVVFTANDSGPR | 101-203 | 613.5567 | 4 | y6+ | 645.2951 |
|  |  |  |  |  | y4+ | 416.2252 |
|  | A**L**[**^13^C_6_**]GISPFHEHAEVVFTANDSGPR |  | 615.0618 | 4 | y6+ | 645.2951 |
|  |  |  |  |  | y4+ | 416.2252 |
|  | TSESGELHGLTTEEEFVEGIYK | 69-90 | 819.0552 | 3 | y4+ | 480.2817 |
|  |  |  |  |  | y5+ | 609.3243 |
|  | TSESGE**L**[**^13^C_6_**]HGLTTEEEFVEGIYK |  | 821.0619 | 3 | y4+ | 480.2817 |
|  |  |  |  |  | y5+ | 609.3243 |
|  | TSESGE**L**[**^13^C_6_**]HG**L**[**^13^C_6_**]TTEEEFVEGIYK |  | 823.0686 | 3 | y4+ | 480.2817 |
|  |  |  |  |  | y5+ | 609.3243 |
| **Internal standard** | | | | | | |
| Yeast enolase | LGANAILGVSLAASR | 106-120 | 706.9146 | 2 | y8+ | 760.4312 |
|  |  |  |  |  | y11+ | 1057.6364 |
|  | GNPTVEVELTTEK | 16-28 | 708.8645 | 2 | y11++ | 623.3323 |
|  |  |  |  |  | y8+ | 948.4884 |

*SILK peptide transitions include heavy isotope labeled leucine and are shown by inclusion of* ***L[^13^C_6_]*** *in the above peptide sequences.*

*Both quantitative and qualitative product ions are listed, with the quantitative ion for each peptide/precursor ion stated first.*
